# Determinants Of Knowledge And Awareness Of Obstetric Danger Signs Among Pregnant Women In Kumasi, Ghana: A Cross-Sectional Study

**DOI:** 10.64898/2026.11.24.26368695

**Authors:** Erica Hammond, Gertrude Oforiwaa Amanfo, Daniella Awere Gambrah, Augustina Kwartemaah Frimpong, Abigail Duku, Veronica .M. Dzomeku, Prince Addo Anim

**Author notes:** Corresponding author: Emory University Hospital, Department of Laboratory Medicine, 1364 Clifton Rd NE, Atlanta, GA 30322, United States of America. *Email address:* (P.A. Anim).

## Abstract

**Background:** Timely recognition of obstetric danger signs during pregnancy, childbirth, and the postpartum period is critical for reducing maternal and neonatal morbidity and mortality. In Ghana, maternal mortality remains high despite improvements in healthcare, partly due to inadequate awareness of these signs. This study assessed the knowledge and awareness of obstetric danger signs among pregnant women attending antenatal care (ANC) at Kwadaso SDA Hospital.

**Methods:** A cross-sectional study was conducted among 200 pregnant women aged 18 years and above, recruited through simple random sampling. Data were collected using structured, interviewer-administered questionnaire adapted from JHPIEGO, covering sociodemographic information, obstetric history, knowledge of danger signs in different pregnancy phases and awareness-related attitudes. Data were analyzed using SPSS version 21, with descriptive statistics, Chi-square tests, and logistic regression used to assess associations. Statistical significance was set at p < 0.05.

**Results:** The most recognized obstetric danger signs were severe vaginal bleeding (97% antepartum, 85% intrapartum, and 92% postpartum) and retained placenta (95% intrapartum). Knowledge was high during the antepartum (86.5%) and intrapartum (78%) periods but dropped to 41% postpartum, resulting in 59% having poor overall knowledge. Women with 1–3 children [(aOR=3.86, 95% CI: 1.31–11.36; p=0.014)] and 4–6 children [(aOR=6.12, 95% CI: 1.43–26.16; p=0.015)] were more likely to have good knowledge. Those with pregnancy-induced hypertension [(aOR=13.25, 95% CI: 1.10–159.02; p=0.042)] and women who attended six to nine ANC visits [(aOR=8.98, 95% CI: 1.44–56.12; p=0.019)] also demonstrated increased likelihood of good knowledge, while participants with only primary education had lower odds [(aOR=0.06, 95% CI: 0.01–0.46; p=0.007)].

**Conclusion:** Although awareness of some key obstetric danger signs, particularly bleeding, was high, overall knowledge remained inadequate, especially in the postpartum period. Strengthening targeted ANC education, with emphasis on postpartum complications, is essential to improve timely care-seeking and maternal outcomes in this population.

## 1. Introduction

Maternal health remains a major problem worldwide, particularly in low- and middle-income countries where maternal mortality and morbidity rates are exceedingly high (Geller et al., 2018). A crucial aspect of reducing maternal mortality is the timely recognition and treatment of obstetric danger signs, which are indicative symptoms that indicate possible complications during pregnancy, childbirth or the postpartum period (Padilla et al., 2021). Timely detection and appropriate interventions can significantly improve outcomes for both mothers and infants (Negero et al., 2021). Despite ongoing efforts to improve maternal health services, there remains a significant lack of understanding and awareness of these warning signs among pregnant women, particularly in resource-limited areas (Subayi, 2023).

Globally, an estimated 295,000 women lost their lives due to pregnancy-related reasons in 2017 alone (Sumankuuro et al., 2022). Most of these fatalities happen in low- and middle-income countries (LMICs), where healthcare systems frequently have difficulty offering sufficient antenatal care (ANC) services (Sumankuuro et al., 2022). In Ghana, despite numerous efforts to improve maternal health, maternal mortality remains alarmingly high. According to the Ghana Health Service (GHS), the country’s maternal mortality rate was estimated to be 319 per 100,000 live births in 2017 (Boafor, 2023). Although this is significantly better than other countries in Sub-Saharan Africa (SSA), it is still below the Sustainable Development Goal (SDG) which aims to reduce maternal mortality to below 70 per 100,000 live births by 2030 (Olea-Ramirez et al., 2024). A major factor contributing to these increased mortality rates is inadequate timely recognition of obstetric complications, which often results in delays in seeking appropriate care or inadequate treatment in emergencies (Akpan et al., 2020).

Obstetric warning signs indicate various symptoms or problems that occur during pregnancy, labor, or the postpartum period and indicate possible life-threatening complications (Gobran et al., 2021). These may include, but are not limited to, severe headaches (Gobran et al., 2021), increased blood pressure (Gobran et al., 2021), vaginal bleeding (Hannola et al., 2021), decreased fetal activity (Heazell et al., 2018), and extreme abdominal discomfort (Huang et al., 2014). Studies have shown that women who are well-informed about these signs are more likely to seek timely medical assistance when complications arise (Asfaha et al., 2022; Phanice & Zachary, 2018; Suglo & Siakwa, 2016; Tamang et al., 2021). Knowing the dangerous signs of pregnancy is a crucial first step in accepting appropriate and timely referrals for obstetric care. It aids in decision-making for seeking medical care in the event of complications: delay in the decision to seek care, delay in reaching the place of care, and delay in receiving appropriate care (Vidler et al., 2023). Knowing the dangerous signs of pregnancy is a crucial first step in accepting appropriate and timely referrals for obstetric care. Furthermore, it aids in decision- making for seeking medical care in the event of complications (Vidler et al., 2023).

Despite the fact that every pregnant mother should recognize obstetric danger signs and symptoms, little is known about this research niche in Ghana. Therefore, the present study aimed to assess the knowledge and awareness of obstetric danger signs among pregnant women attending antenatal care at Kwadaso SDA Hospital.

## 2. Methods

### 2.1 Study designs and Demography

This was a cross-sectional study conducted among pregnant women at the Kwadaso SDA Hospital. The Hospital has a 150-bed capacity and about 50 doctors in various specialties. The facility also features a robust ANC Clinic that caters for a diverse population of pregnant women.

This facility was chosen due to its strategic location and the volume of antenatal care services it provides. The hospital’s robust ANC Clinic is well-equipped to handle a large number of patients, offering comprehensive maternal health services. This made it an ideal setting for evaluating the knowledge and awareness of obstetric danger signs among pregnant women (KwadasoSDAHospital, 2024).

### 2.2 Study population

The sample size was determined using the Yamane formula: 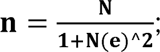

where n = Sample size, N = population size= estimated population of pregnant women

Estimated number of pregnant women utilizing ANC at the Kwadaso SDA Hospital is 300. Therefore, N = 300, e = confidence level/ margin of error (0.05).

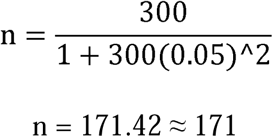

The minimum sample required for the study is one hundred and seventy -one (171). However, 200 participants were recruited for this study in order to increase statistical power.

### 2.3 Inclusion criteria

This study included pregnant women who were aged 18 years and above, attending Antenatal Care at the Kwadaso SDA Hospital and willing to participate in the study.

### 2.4 Exclusion Criteria

This study excluded non-pregnant women, pregnant women below 18 years and women with severe cognitive impairment.

### 2.5 Data Collection

Data was collected through face-to-face interviews using pre-tested structured questionnaire adapted from the Maternal and Neonatal Program of JHPIEGO, an affiliate of John Hopkins University (JHPIEGO, 2004). The questionnaire collected demographic information, details about the participant’s obstetric history, and their knowledge of key danger signs during pregnancy, childbirth, and the post-partum period. The questionnaire also evaluated the participants’ awareness of the importance of recognizing these signs and seeking prompt medical attention. Participants were deemed to have good knowledge if they can spontaneously mention at least three key danger signs from each phase of pregnancy, childbirth, and the post-partum period. Overall good knowledge was defined as participants demonstrating good knowledge across all three phases of pregnancy. Participants’ written consent was sought after which trained research assistants administered the questionnaire, sometimes translating into the local languages that the respondents could understand.

### 2.6 Statistical Analysis

The data collected was entered into Microsoft Excel and screened for errors. Subsequently, the data was analyzed using the Statistical Package for the Social Sciences (SPSS) version 21 (SPSS Inc., Chicago, Ill., USA) and Graphpad Prism (www.graphpad.com). Categorical variables were summarized as frequencies and percentages. Chi-square tests and Binary logistic regression was used to determine associations between study variables and good knowledge of obstetric danger signs among pregnant women. 95% confidence interval and p-value less than 0.05 were considered statistically significant.

## 3.0 Results

### 3.1 Sociodemographic characteristics of study participants

Out of the 200 participants, most were aged 26–30 years (35.5%), followed by 31–45 years (34.0%) and 18–25 years (30.5%). The majority were married (59.5%), while 35.0% were single. Tertiary education was most common among participants (43.5%), followed by secondary education (38.5%), with only 8.0% having no formal education. Over half of the participants’ partners had tertiary education (56.0%), and 34.5% had secondary education. Most participants were either self-employed (25.5%) or unemployed (25.0%). Household income ranged mostly between GHS 2000–2999 (36.0%) and ≥GHS 3000 (30.5%). A majority had 1–3 children (57.5%), and 29.0% had no children. Most identified as Christians (78.5%) and lived in urban areas (99.0%). **(Table 1).**

**Table 1:**
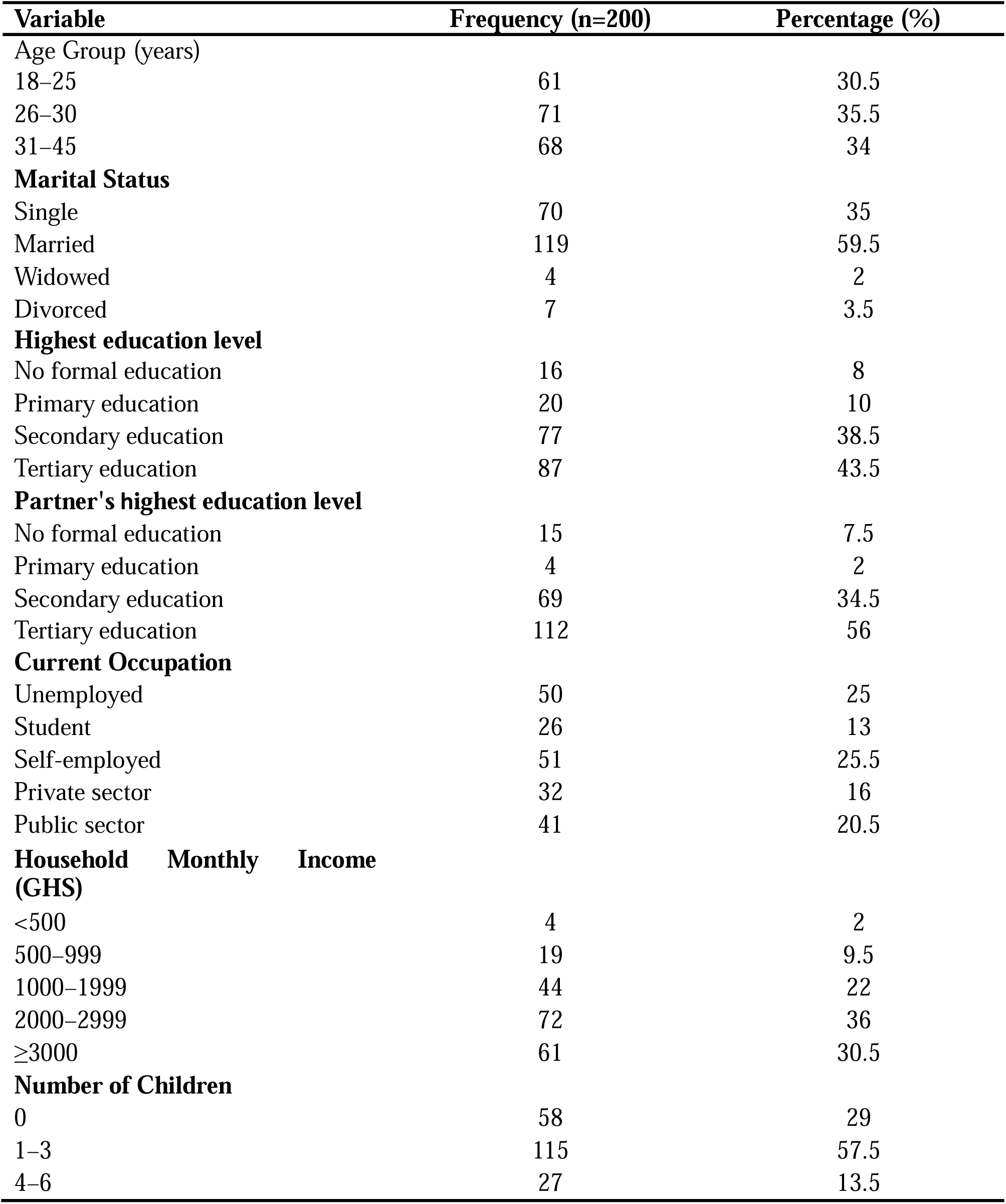
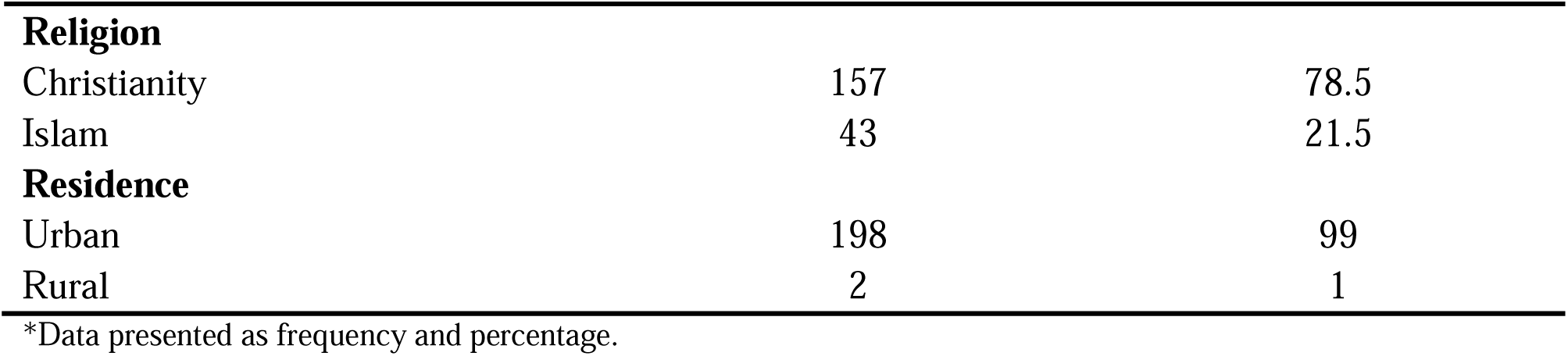
Sociodemographic Characteristics of study participants.

#### 3.2.1 Obstetric Characteristics of study participants

Most participants had been pregnant 1–4 times (62.0%), while 22.5% were in their first pregnancy. The majority were in their second trimester (54.5%), followed by the first (26.0%) and third (19.5%) trimesters. Regarding previous life births, most had 1–3 previous births (59.0%), while 28.5% had none. A majority reported no complications in previous pregnancies (75.5%), though some experienced miscarriage (12.0%), preeclampsia (5.0%), or preterm labor (4.5%). Other less common complications included gestational diabetes (4.0%) and abortion (2.5%). In the current pregnancy, most had no complications (87.0%). Minor complications reported included pregnancy-induced hypertension (3.0%), swollen extremities (2.5%), abdominal pain (1.5%), vaginal discharge with odor or itching (1.0%), and gestational diabetes (1.5%). Most participants had visited the antenatal clinic 1–3 times (46.5%) or 4–6 times (45.5%), while 8.0% had 7–9 visits **(Table 2).**

**Table 2:**
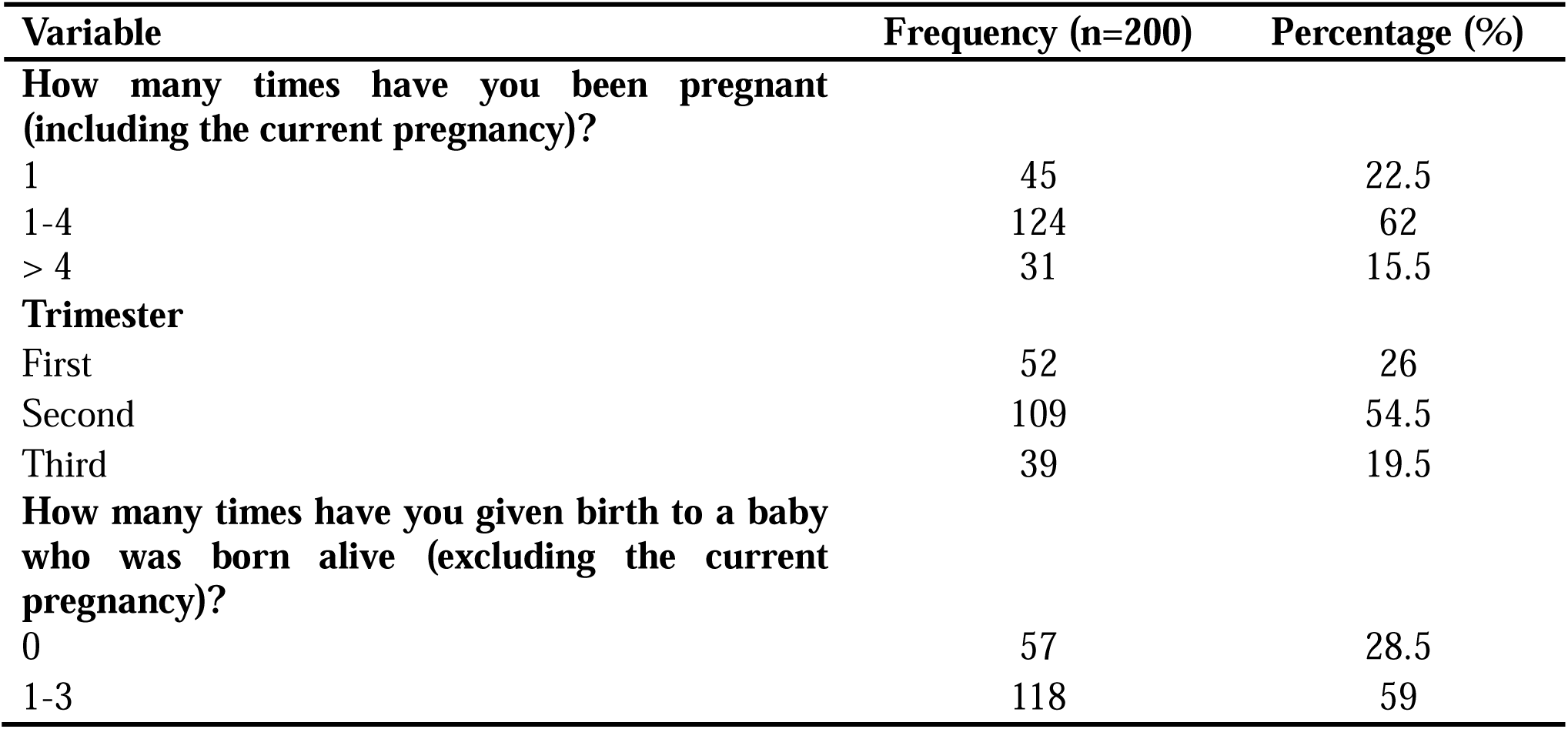
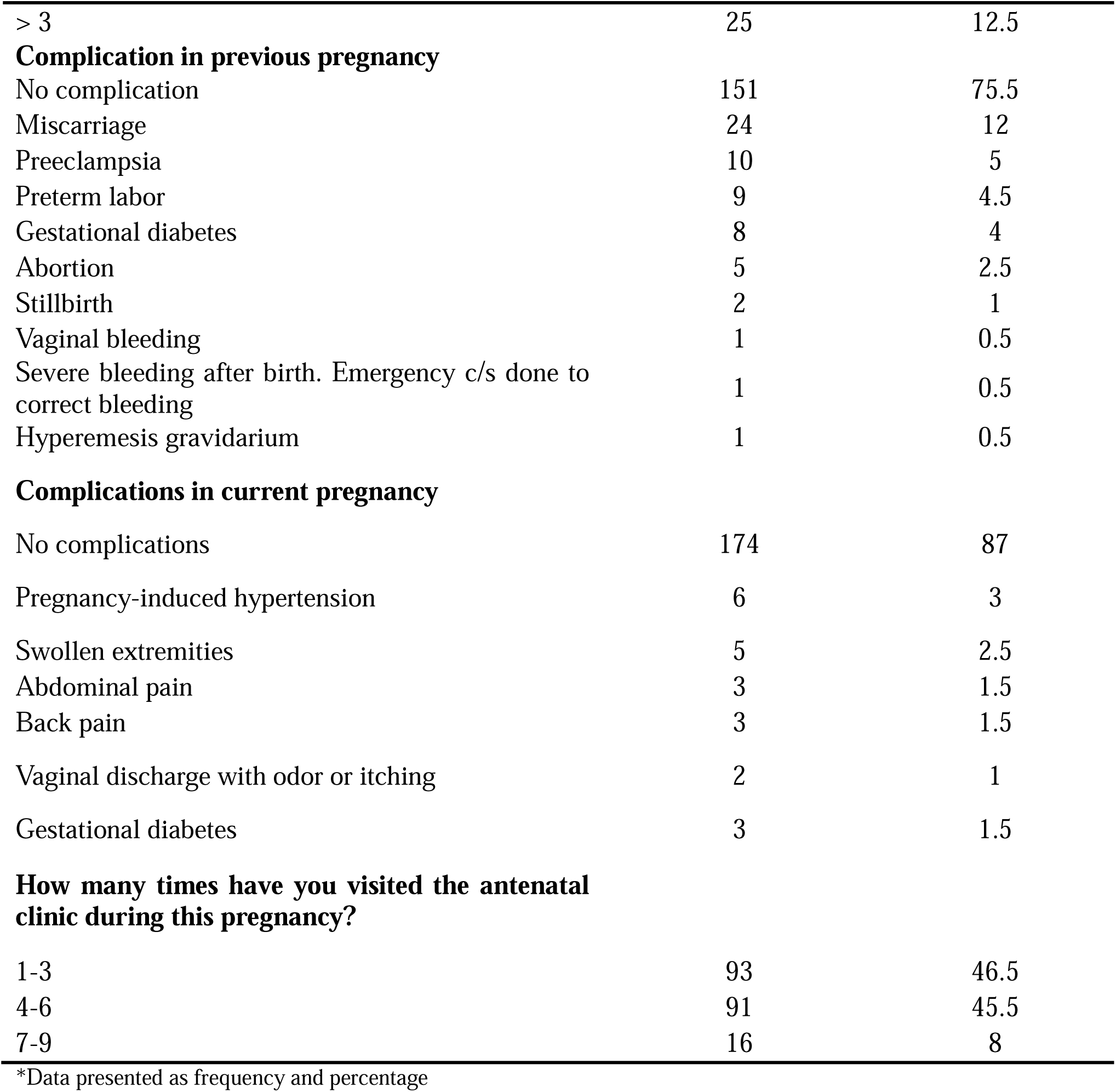
Obstetric Characteristics of study participants.

#### 3.2.2 Obstetric danger signs identified in the antepartum phase

The most commonly identified danger signs in the antepartum period were severe vaginal bleeding (97.0%), followed by no or reduced fetal movement (81.0%), water breaking without labor (77.0%), and severe pelvic or abdominal pain (71.5%). Over half of the participants mentioned severe headache (57.0%), swollen hands, face, and/or feet (54.5%), and loss of consciousness (52.0%). Convulsions (30.5%), blurred vision (35.5%), hyperemesis gravidarum (1.0%), and high fever (0.5%) were less frequently reported (Table 3).

**Table 3:**
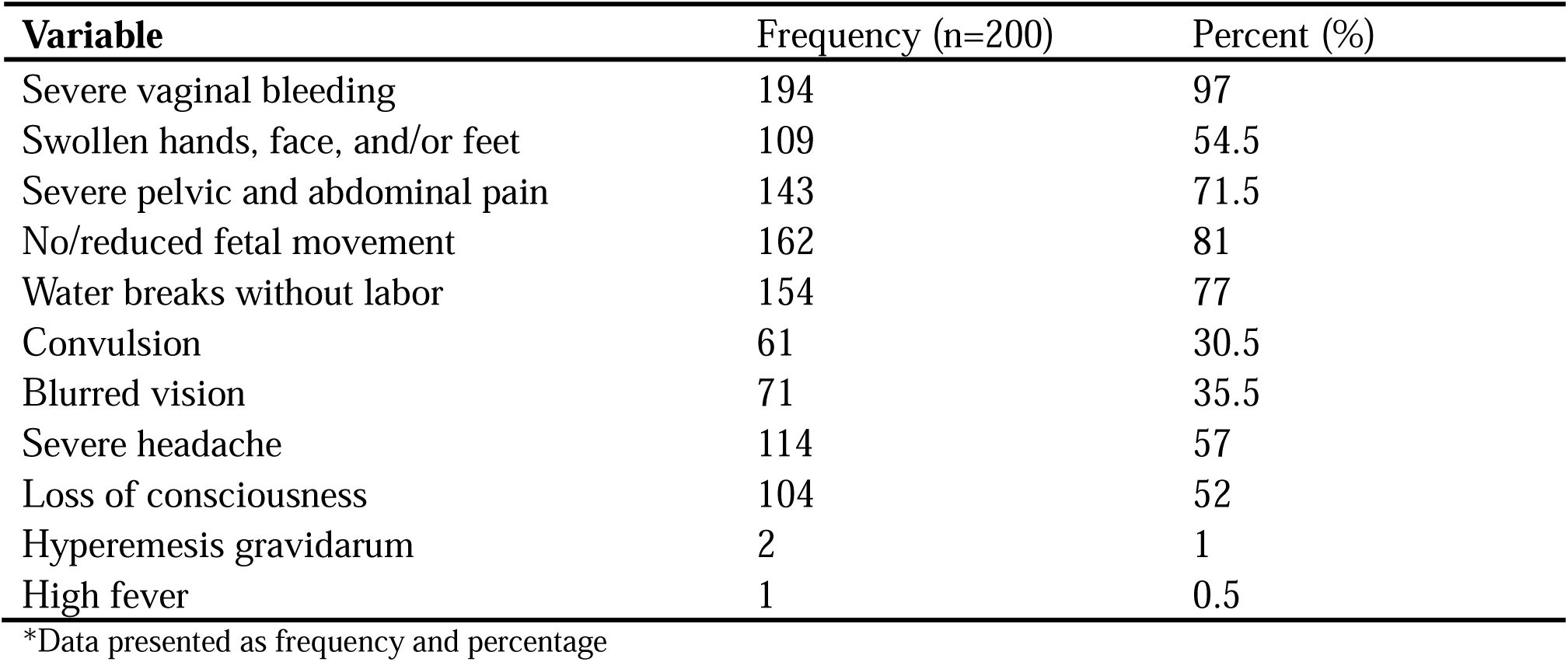
Obstetric danger signs identified in the antepartum phase.

#### 3.2.3 Obstetric danger signs identified in the intrapartum phase

The most frequently identified intrapartum danger signs were retained placenta (95.0%) and severe vaginal bleeding (85.0%). Labor lasting more than 12 hours (68.0%), cord presenting before the baby (57.0%), and limb presentation (55.5%) were also commonly mentioned. Less frequently identified signs included loss of consciousness (39.0%), difficulty breathing (25.0%), and convulsions (15.5%) **(Table 4).**

**Table 4:**
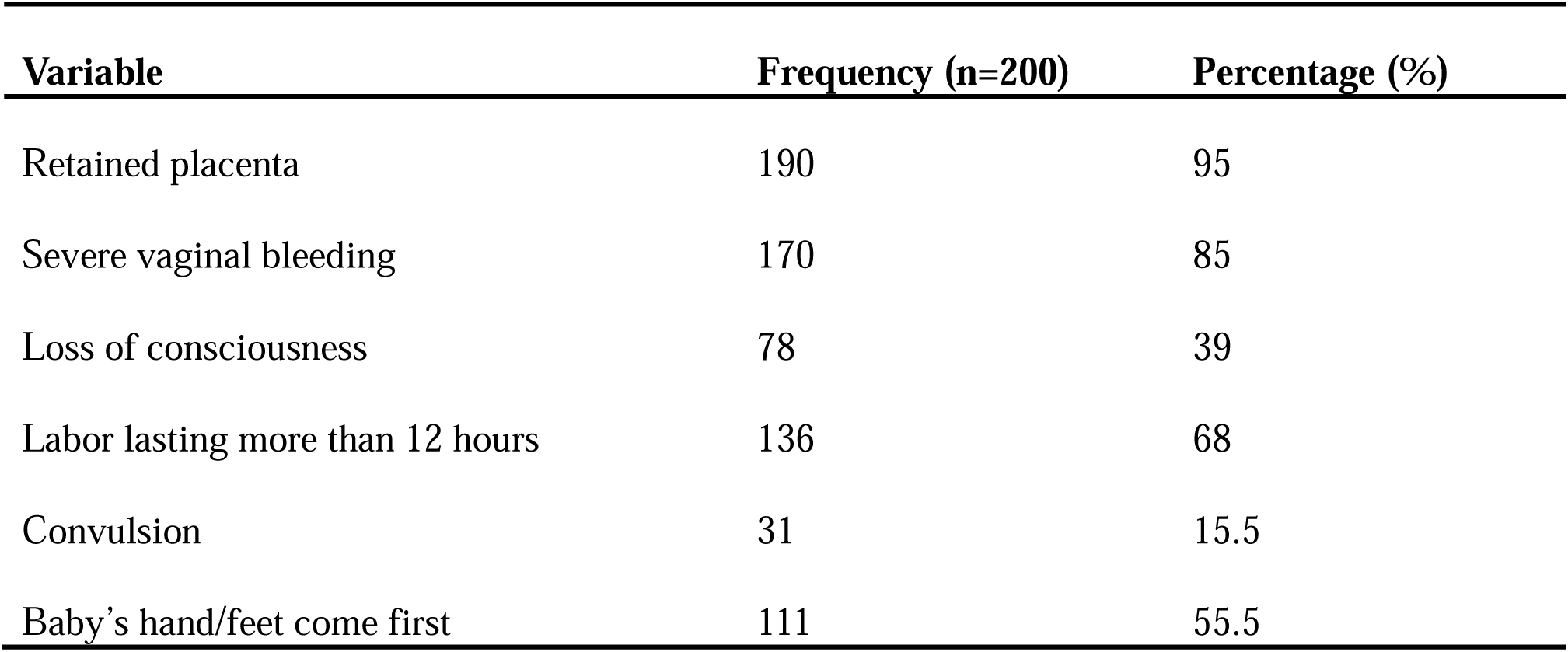
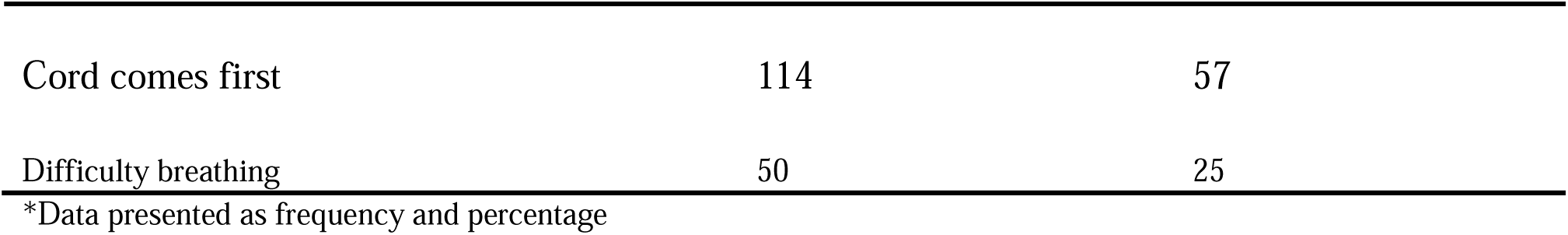
Obstetric danger signs identified in the intrapartum phase.

#### 3.2.4 Obstetric danger signs identified in the postpartum phase

Severe vaginal bleeding (92%) was the most frequently identified obstetric danger sign in the postpartum phase. High fever (42.5%) and swollen hands, face, and/or feet (36.5%) were also commonly recognized. Foul-smelling lochia (34%) was reported by over one-third of the participants. Difficulty breathing (9.5%), convulsion (3.5%), severe lower abdominal pain (1%), and loss of consciousness (0.5%) were identified less frequently **(Table 5)**

**Table 5:**
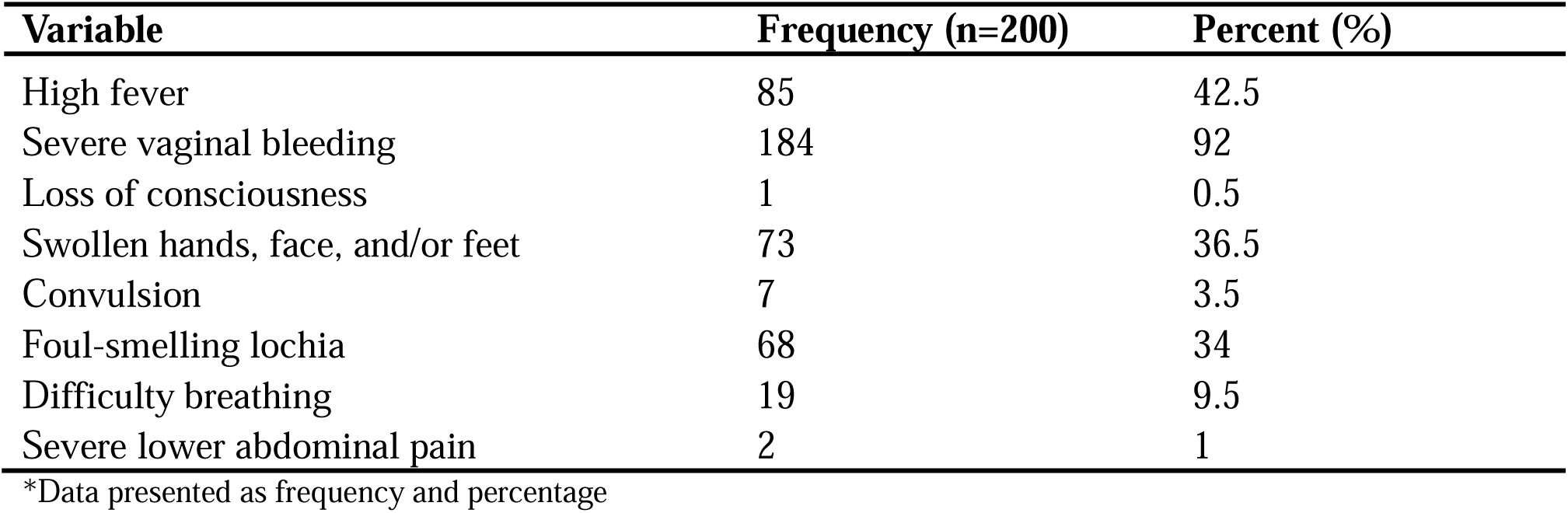
Obstetric danger signs identified in the postpartum phase.

### 3.3 Knowledge level of obstetric danger signs across various phases of pregnancy

Knowledge of obstetric danger signs varied across the perinatal periods, with most participants demonstrating good knowledge during the antepartum phase (86.5%). This level of understanding decreased somewhat in the intrapartum period, where a sizable majority still showed good knowledge (78%). However, knowledge significantly declined in the postpartum period, with less than half of participants demonstrating adequate awareness (41%). Consequently, overall knowledge of obstetric danger signs was poor for the majority of participants (59%) **(Figure 1)**

**Figure 1:**
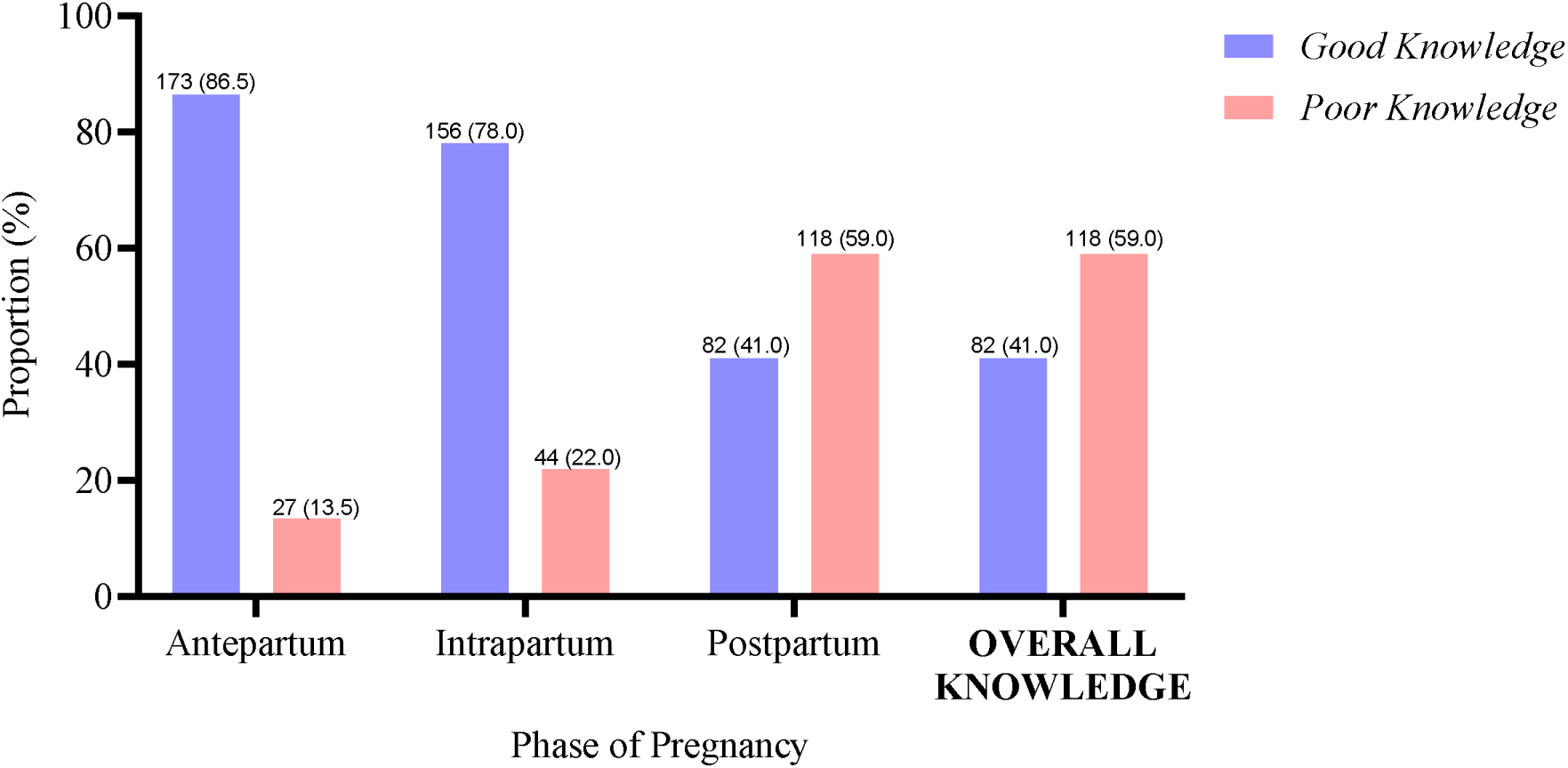
Knowledge level of obstetric danger signs across various phases of pregnancy.

**Figure 1:**
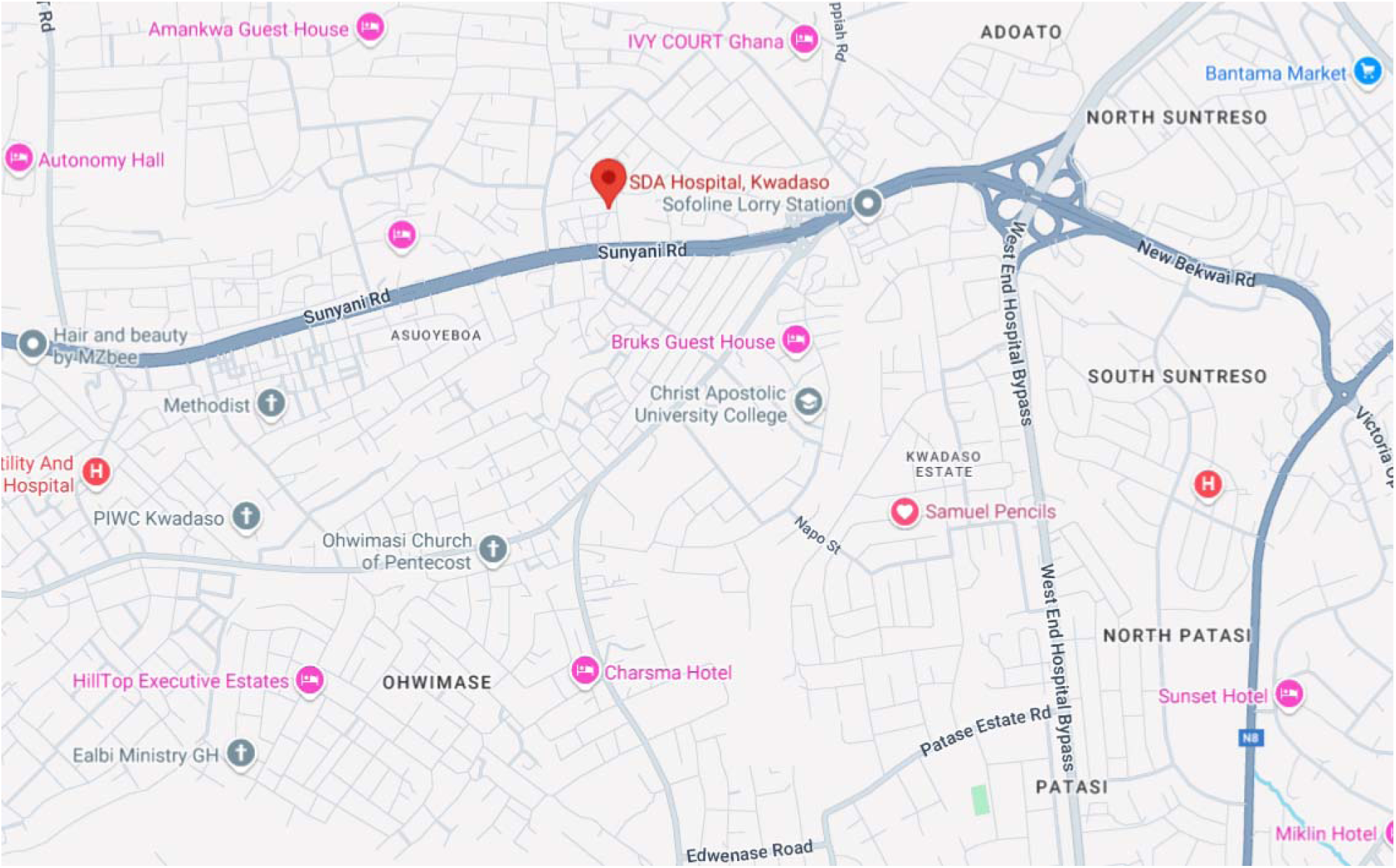
Location of Kwadaso SDA Hospital.

### 3.4 Association between sociodemographic variables and good knowledge of obstetric danger signs

Age group (p < 0.001), marital status (p = 0.014), educational level (p = 0.001), occupation (p = 0.016), monthly household income (p = 0.001), and number of children (p < 0.001) showed statistically significant associations with knowledge of obstetric danger signs. In contrast, partner’s educational level (p = 0.888), religion (p = 0.407), residence (p = 0.795), and ethnicity (p = 0.176) did not exhibit significant associations (Table 6).

**Table 6:**
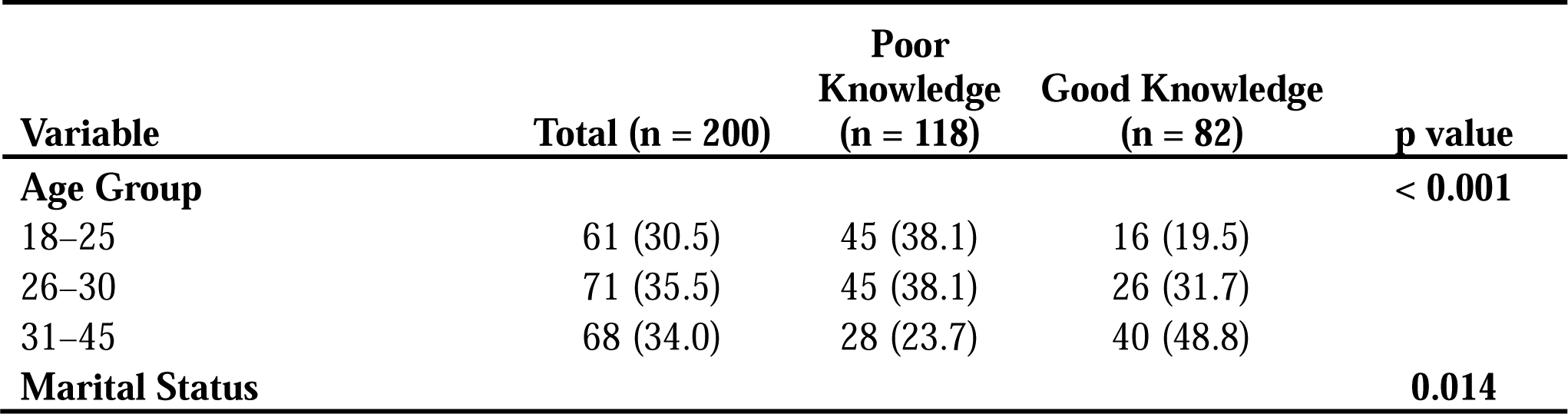
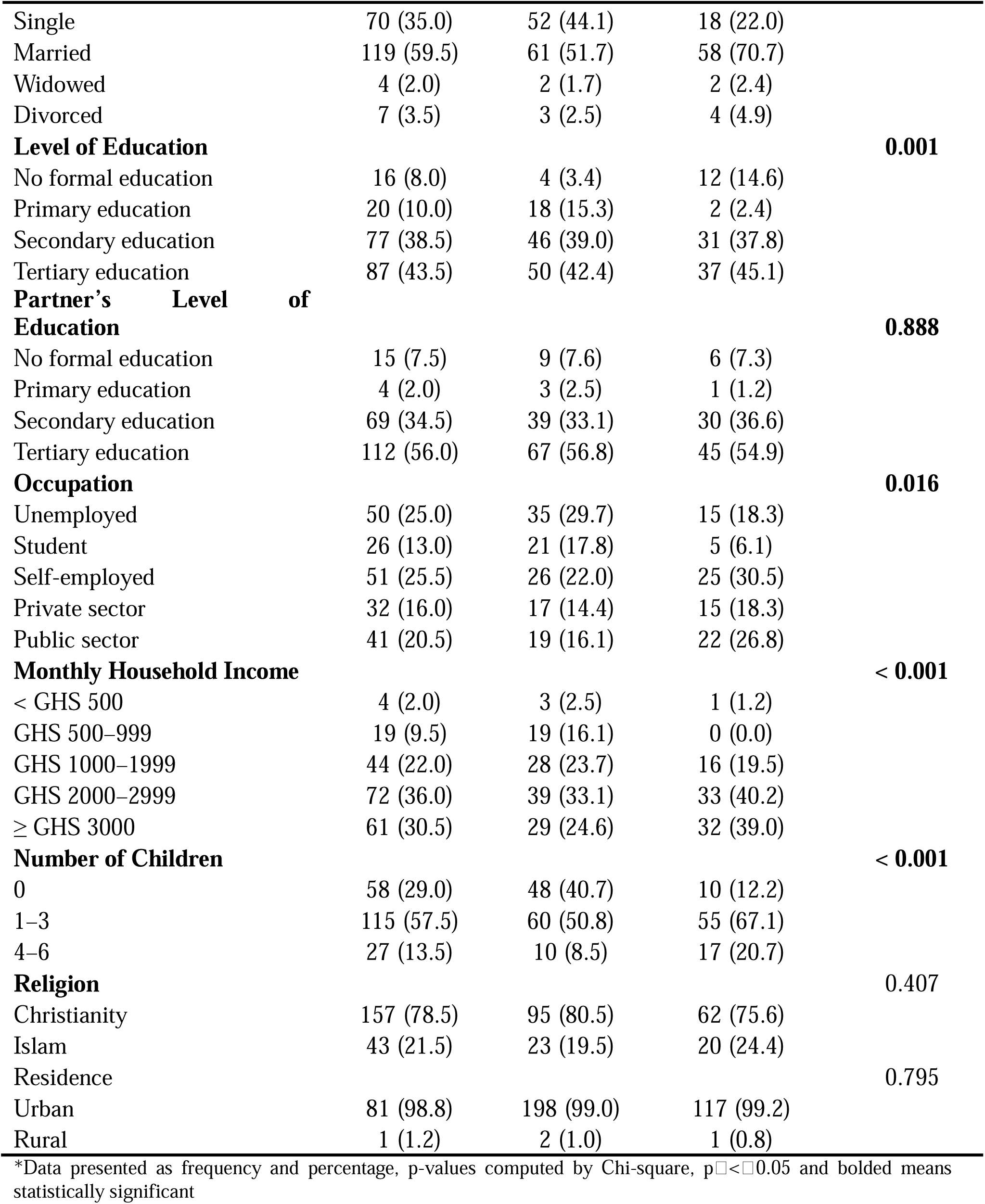
Association between sociodemographic variables and good knowledge of obstetric danger signs.

### 3.5 Association between sociodemographic variables and good knowledge of obstetric danger signs

Gravidity (p < 0.001), trimester of pregnancy (p = 0.002), parity (p < 0.001), history of complications in previous pregnancies (p = 0.001), complications in current pregnancy (p = 0.001), and number of antenatal care visits during the current pregnancy (p = 0.004) showed statistically significant associations with knowledge of good knowledge of obstetric danger signs **(Table 7)**.

**Table 7:**
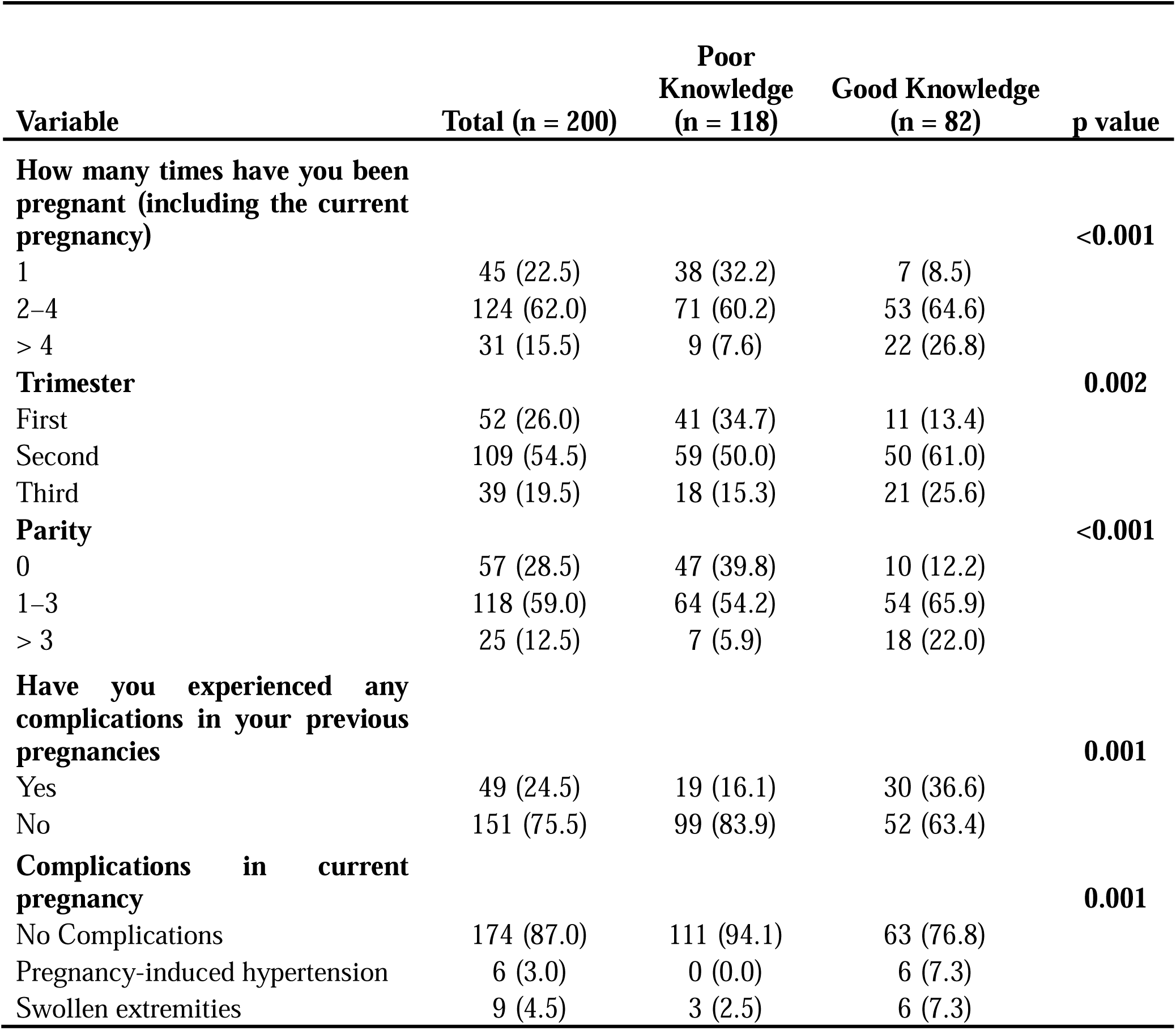
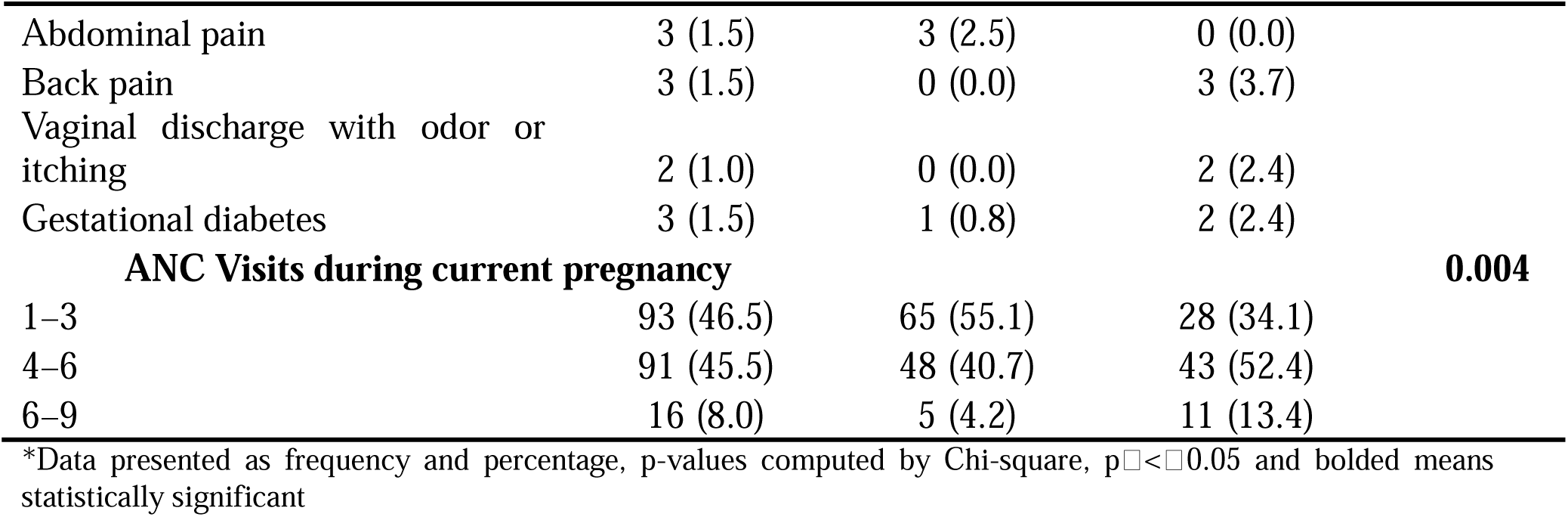
Association between sociodemographic variables and good knowledge of obstetric danger signs.

### 3.6 Sociodemographic Predictors of good knowledge of obstetric danger signs

In a univariate logistic regression model, participants aged 31–45 years were four times more likely to have good knowledge of obstetric danger signs compared to those aged 18–25 years [cOR = 4.02, 95% CI (1.90–8.48), p < 0.001]. Married participants were 2.75 times more likely than single participants to have good knowledge [cOR = 2.75, 95% CI (1.44–5.24), p = 0.002]. Employment in the public sector was associated with 2.70 times increased odds of good knowledge compared to being unemployed [cOR = 2.70, 95% CI (1.14–6.40), p = 0.024].

Unexpectedly, having any level of formal education was associated with reduced odds of good knowledge when compared to no formal education: primary education [cOR = 0.04, 95% CI (0.006–0.24), p < 0.001], secondary education [cOR = 0.23, 95% CI (0.066–0.76), p = 0.016], and tertiary education [cOR = 0.25, 95% CI (0.074–0.83), p = 0.023]. Participants with 1–3 children had 4.40 times higher odds of good knowledge [cOR = 4.40, 95% CI (2.03–9.54), p < 0.001], while those with 4–6 children had 8.16 times higher odds [cOR = 8.16, 95% CI (2.89–23.01), p < 0.001], compared to those without children.

In a multivariate logistic regression model, only the number of children remained significantly associated with good knowledge. Participants with 1–3 children had 3.86 times higher odds of good knowledge compared to those without children [aOR = 3.86, 95% CI (1.31–11.36), p = 0.014], and those with 4–6 children had 6.12 times higher odds [aOR = 6.12, 95% CI (1.43–26.16), p = 0.015]. Additionally, individuals with primary education had significantly lower odds of good knowledge compared to those with no formal education [aOR = 0.06, 95% CI (0.01–0.46), p = 0.007]. Other variables, including age, marital status, occupation, and income, were not significantly associated with good knowledge after adjustment. **(Table 8)**.

**Table 8:**
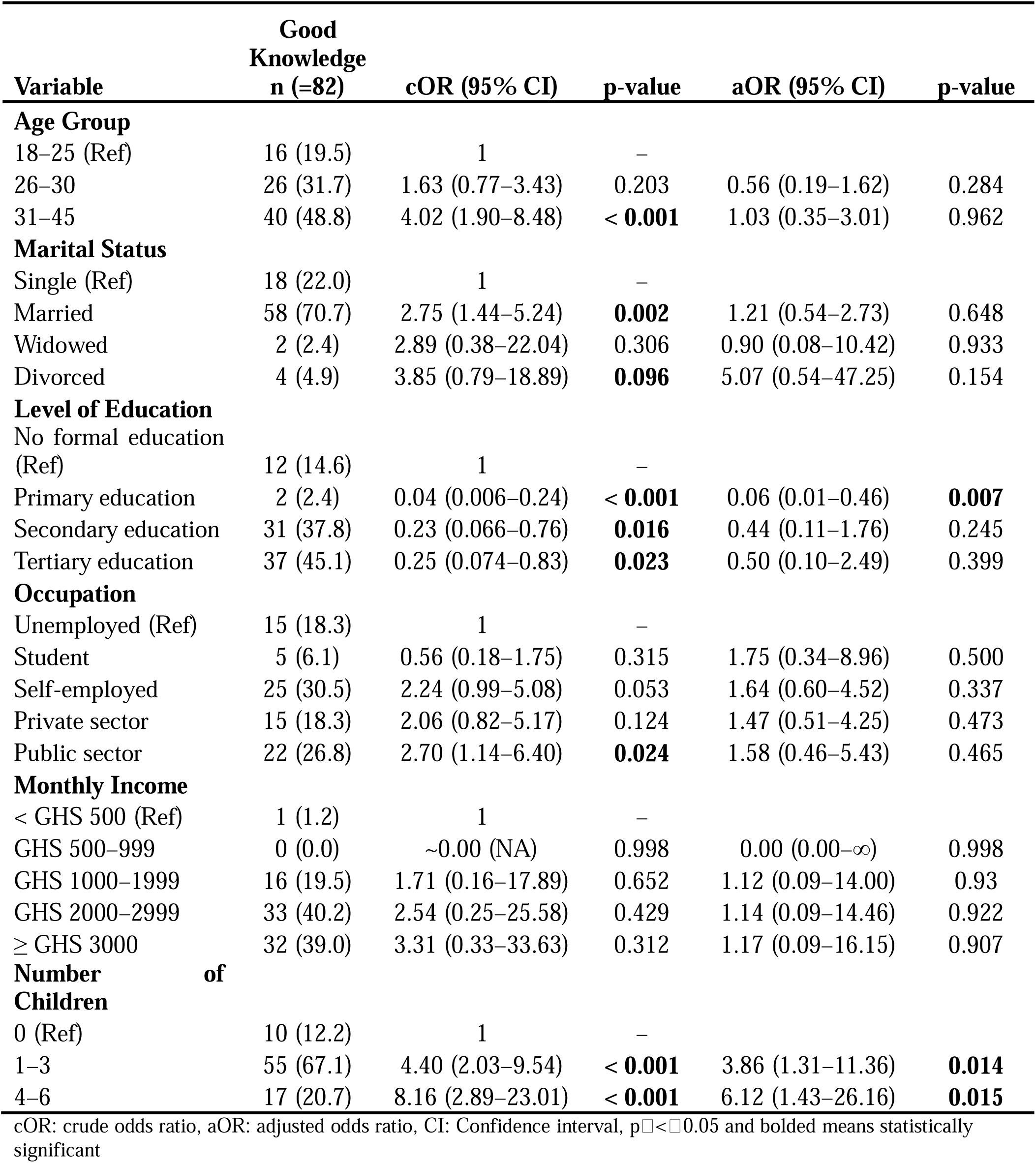
Sociodemographic Predictors of good knowledge of obstetric danger signs.

### 3.7 Obstetric Predictors of good knowledge of obstetric danger signs

In univariate logistic regression, several factors significantly influenced the likelihood of good knowledge of obstetric danger signs. Women with 2–4 pregnancies had a 4.05-fold increased likelihood (cOR = 4.05, 95% CI [1.68–9.78], p = 0.002), and those with >4 pregnancies had a 13.27-fold increase (cOR = 13.27, 95% CI [4.34–40.61], p < 0.001) compared to one pregnancy. Being in the second and third trimesters increased the likelihood by 3.16-fold (cOR = 3.16, 95% CI [1.47–6.79], p = 0.003) and 4.35-fold (cOR = 4.35, 95% CI [1.74–10.87], p = 0.002), respectively, relative to the first trimester. Parity of 1–3 and >3 raised the odds by 3.97-fold (cOR = 3.97, 95% CI [1.83–8.59], p < 0.001) and 12.09-fold (cOR = 12.09, 95% CI [3.99–36.61], p < 0.001) compared to nulliparity. Absence of previous pregnancy complications reduced the likelihood by 67% (cOR = 0.33, 95% CI [0.17–0.65], p = 0.001). Pregnancy-induced hypertension and gestational diabetes showed elevated odds (7.05- and 3.52-fold) but were not statistically significant. Attending 4–6 and 6–9 antenatal visits increased the likelihood by 2.08- fold (cOR = 2.08, 95% CI [1.14–3.81], p = 0.018) and 5.11-fold (cOR = 5.11, 95% CI [1.62–16.07], p = 0.005), respectively, compared to 1–3 visits.

After adjustment, ‘participants with no complication in previous pregnancies had about 80%’ reduced likelihood of having good knowledge of obstetric danger signs (aOR = 0.24, 95% CI [0.09–0.65}. p = 0.005). Conversely, pregnant women who have experienced pregnancy induced hypertension had a 13times increased odds of having good knowledge of obstetric danger signs compared to those who have experienced no complication (aOR = 13.25, 95% CI [1.10–159.02]. p = 0.042). Similarly, participants who has attended ANC between six to nine times had about a nine-fold increased chance of good knowledge of obstetric danger signs (Table 9).

**Table 9:**
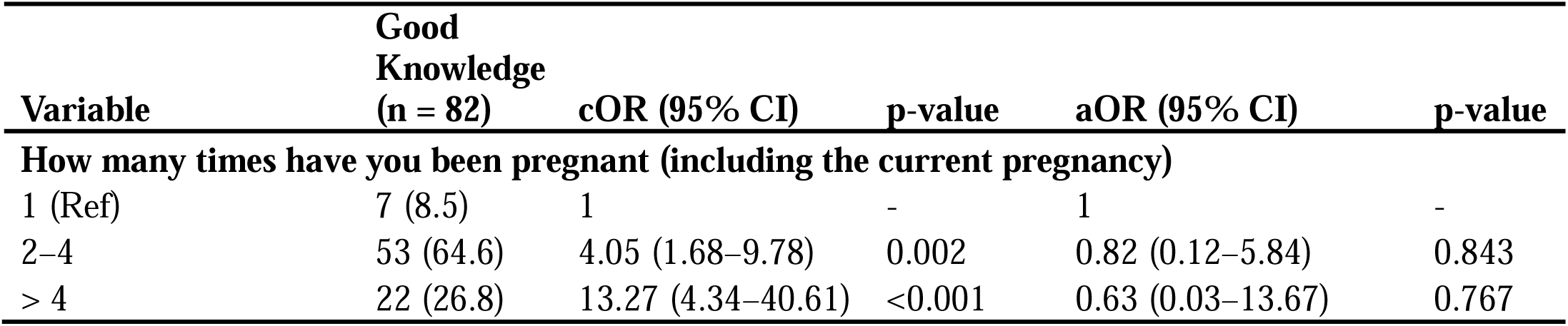
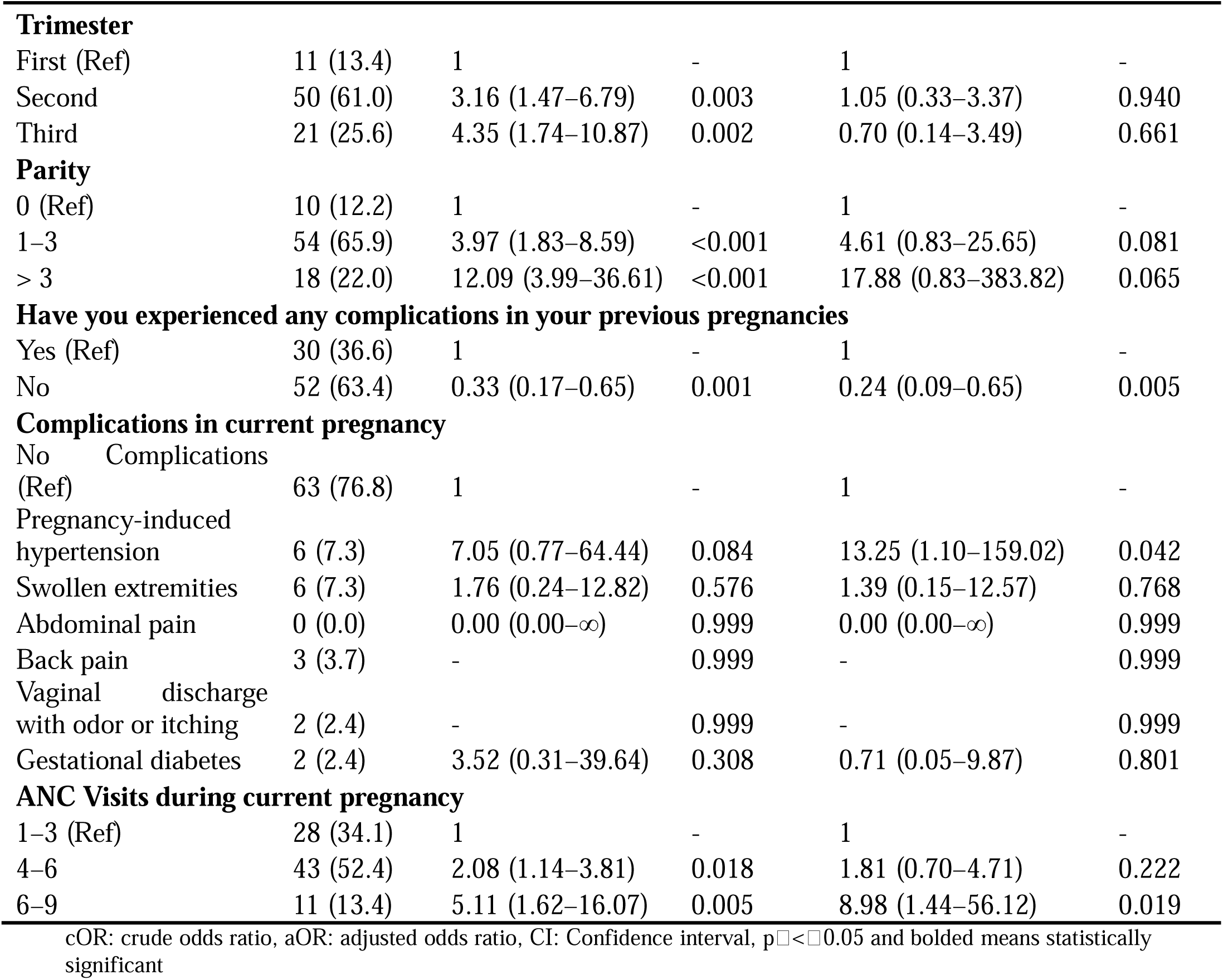
Obstetric Predictors of good knowledge of obstetric danger signs.

### 3.8 Awareness of Obstetric danger signs

The majority (90.5%) recognized the importance of having an emergency plan during pregnancy, with 181 women agreeing. Similarly, 86.5% (n = 173) acknowledged that recognizing warning signs is critical for both maternal and fetal health. Awareness that danger signs require immediate medical attention was reported by 79.0% (n = 158), while 77.0% (n = 154) agreed that seeking help for serious symptoms can protect their health. Additionally, 76.5% (n = 153) recognized that early detection and treatment of complications improve delivery outcomes, and 73.5% (n = 147) understood that awareness can help prevent serious complications or death **(Table 10)**.

**Table 10:**
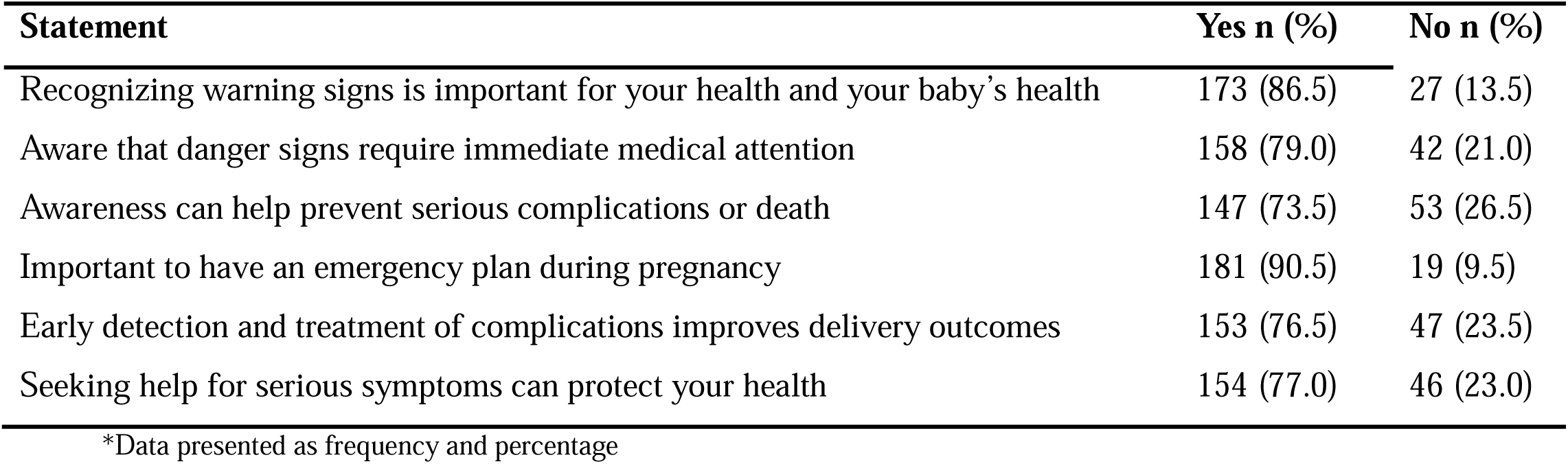
Awareness of Obstetric danger signs.

## 4.0 Discussion

Knowledge of obstetric danger signs is a critical component of maternal health, as it influences how quickly pregnant women recognize complications and seek timely medical attention. Adequate awareness can prevent delays in care, reduce maternal morbidity and mortality, and improve pregnancy outcomes. In this study of 200 pregnant women attending ANC at Kwadaso SDA Hospital, awareness of obstetric danger signs was highest for severe vaginal bleeding across all perinatal periods: 97% antepartum, 85% intrapartum, and 92% postpartum.

In this study, majority of participants (97.0%) identified severe vaginal bleeding in pregnancy as a danger sign, followed by reduced fetal movement (81.0%) and premature rupture of membranes (“water breaking” without labor) (77.0%). These high percentages exceed figures reported elsewhere in Africa. For example, only 45.9% of 744 Ethiopian women in the Aleta Wondo study mentioned antepartum bleeding (Hailu et al., 2010), and 70.9% of 382 Rwandan women did so (Uwiringiyimana et al., 2022), both substantially lower than our 97%. Similarly, awareness of other antepartum signs in our study (e.g. severe abdominal pain 71.5%, swelling of hands/face 54.5%, headache 57.0%, blurred vision 35.5%, convulsions 30.5%) generally exceeded that seen in other studies (Hailu et al., 2010; Uwiringiyimana et al., 2022). In Ghana, a recent postpartum survey likewise found very high awareness of hemorrhage: 94.7% of 151 women at Tamale West Hospital identified severe bleeding (Lau et al., 2023). By contrast, more subtle signs were less recognized. For instance, only about 30–36% of participants in this study knew about convulsions or blurred vision antepartum, reflecting the global pattern that classic danger signs like bleeding are better known than others (Hailu et al., 2010; Uwiringiyimana et al., 2022).

During labor and delivery, retained placenta was identified by 95.0% of our Ghanaian respondents, and severe vaginal bleeding by 85.0%. The majority (68.0%) also named labor lasting greater than 12 hours, cord prolapse (57.0%), and breech presentation (55.5%). These rates are comparable to high levels seen in other settings. For example, a Kenyan study of 275 nomadic women in Pokot found 92.4% citing postpartum hemorrhage, 85.8% citing prolonged labor, and 75.6% citing retained placenta (Kiptulon et al., 2025). In that study, 63.3% knew convulsions were a danger sign and 49.8% mentioned severe headache, again higher than our study figures of 15.5% (convulsions) and 35.5% (headache). Similarly, Ethiopian women in Sidama Zone recognized bleeding (55.0%) and retained placenta (51.4%) during childbirth (Hailu et al., 2010), well below the levels identified in our study in Ghana.

In the early postpartum phase, severe vaginal bleeding was again the most recognized danger sign (92.0% in our Ghana sample). High fever (42.5%), swelling of hands/face (36.5%), and foul-smelling lochia (bad-smelling discharge) (34.0%) were mentioned by a substantial minority. These findings align in part with other African studies. For instance, a Ghanaian survey of 151 women in Tamale found 94.7% identified heavy bleeding and 82.1% identified fever after birth (Lau et al., 2023). The postpartum hemorrhage awareness in our study (92%) matches that Ghanaian Study and far exceeds some other contexts (e.g. only 59% of Ethiopian women (Hailu, 2010 #469) and 50.6% of Nigerian women (Oni et al., 2025) identified postpartum bleeding). Notably, we found higher awareness of malodorous lochia (34%) than the 11.3% seen in the Ethiopian cohort, although lower than the 55.6% in the Nigerian survey (Oni et al., 2025). This adds to the consistent trend that hemorrhage tends to be known as a danger sign by most women, whereas neurological or infective signs are less often recognized Knowledge of obstetric danger signs differed markedly by perinatal period in this study. During pregnancy (antepartum) most women demonstrated good awareness; 86.5% could correctly identify key danger signs. This high antepartum knowledge likely reflects the emphasis on pregnancy-related risks during antenatal education sessions. Knowledge remained relatively high during labor and delivery (intrapartum), with 78% of women still showing good understanding of danger signs at that stage. However, after childbirth the picture changed dramatically: only 41% of women demonstrated adequate awareness of postpartum danger signs. Thus, the overall level of knowledge across all stages was poor for the majority of participants (59% had inadequate overall knowledge).

The steep decline in awareness after delivery is concerning because the postpartum period carries significant risks for new mothers. Complications such as hemorrhage and infection often arise in the days and weeks after birth, so women need to recognize warning signs to seek timely care (Lau et al., 2023). The relatively low postpartum knowledge observed here suggests that counseling about post-birth danger signs may be insufficient or less well retained. This gap has been noted elsewhere; in Ghana, for example, Lau et al. emphasize that all women need comprehensive discharge education on post-birth warning signs, because promoting this knowledge “can decrease delays in seeking care” and help reduce maternal mortality (Lau et al., 2023). Inadequate postpartum education may therefore contribute to delayed care-seeking in this setting.

Despite strong antenatal awareness, the finding that 59% of women had poor overall knowledge of obstetric danger signs highlights a general need for improved health education. Low awareness of danger signs is common in resource-limited settings, and it directly undermines timely care-seeking. For instance, a recent study in the Democratic Republic of Congo found that pregnant women’s knowledge of obstetric danger signs was very low (21.9%), warning that without better awareness it is “difficult for women to take prompt decisions to seek emergency obstetric care” (Imani Ramazani et al., 2023). Likewise, an Ethiopian study noted that increasing knowledge of key danger signs – through both facility-based and community education – is essential to “make women and their families ready for prompt and appropriate decisions” when complications occur (Hibstu & Siyoum, 2017)

In the present study, the only demographic factor independently linked to better danger sign knowledge was parity (number of children). Women with 1–3 children had about 3.9× higher odds (aOR≈3.86) and those with 4–6 children about 6.1× higher odds (aOR≈6.12) of good knowledge than nulliparous women. This suggests that experience from previous births or pregnancies enhances awareness of warning signs. Similar patterns appear in other settings. For example, a nationwide survey in the Democratic Republic of Congo found that multigravida women listed more danger signs than primigravida’s (Nkamba et al., 2021). In Ghana, Peprah et al. (2023) studied 399 pregnant women (Upper West Region) and likewise reported that women with prior deliveries were much less likely to have poor danger sign knowledge than first-time mothers (Peprah et al., 2025).

Education showed an unexpected association. Women with only primary schooling had far lower odds of good knowledge (aOR≈0.06) than women with no formal education. This contradicts the usual finding that higher education improves awareness. For instance, a facility-based study in Ethiopia found that mothers with primary or higher education were 4.85–6.90 times more likely to know pregnancy danger signs than uneducated mothers (Gesese et al., 2023). Likewise, a Ghanaian study of postpartum women with preeclampsia (Korle Bu, Accra; n=150) reported that higher educational level and more antenatal counselling were associated with better knowledge scores (Joshi et al., 2020). In contrast, Peprah et al. found no significant bivariate association between maternal education and danger sign knowledge (Peprah et al., 2025). The discrepancy suggests that in our context, having only basic education may not suffice; it may reflect that formal schooling per se does not guarantee health knowledge unless accompanied by effective antenatal education or higher literacy.

Furthermore, ANC attendance also had a strong effect. Mothers with 6–9 ANC visits had about 9× higher odds of good knowledge than those with fewer visits. This aligns with other evidence that repeated ANC contacts build knowledge. For example, an Ethiopian ANC study (n=414) reported that women with *any* prior ANC visit were 9.27 times more likely to know danger signs than women with no previous visits (Gesese et al., 2023). Similarly, the DRC analysis found that having more ANC visits significantly increased the number of danger signs women could recall (Nkamba et al., 2021). Each ANC visit provides additional health education; the more visits a woman attends, the more exposure she has to counseling on obstetric warning signs. This cumulative effect is consistent across settings: repeated antenatal counseling is a well-recognized driver of maternal knowledge (Afaya et al., 2020).

## 5.0 Conclusion

This study highlights that knowledge of obstetric danger signs among pregnant women in Ghana is unevenly distributed, with certain experiential and clinical factors exerting a strong influence. Women with more children, prior pregnancy complications, or greater ANC engagement demonstrated enhanced awareness, suggesting that repeated exposure to maternity services and firsthand experiences of complications can foster better recognition of danger signs. Conversely, limited education and lack of pregnancy-related challenges appeared to reduce awareness, underscoring the role of both formal learning and lived maternal experiences in shaping knowledge. These findings point to the fact that the determinants of maternal health literacy are not purely informational but are intertwined with socio-demographic, obstetric, and health system factors.

## Supporting information

supplemental list of tables

## Data Availability

All data produced in the present study are available upon reasonable request to the authors

## Abbreviations

ANC: Antenatal Care
CHRPE: Committee on Human Research, Publication and Ethics
KNUST: Kwame Nkrumah University of Science of Technology

## 6.0 Acknowledgement

The authors are grateful to study participants, as well as research assistants who contributed in diverse ways to the successful implementation of the study.

## 7.0 Author contributions: CRediT

EH, GOA, PAA and VMD conceptualized the study with supporting input from AKF. EH and PAA led methodology, data curation, formal analysis and investigation, with DAG and GOA supporting screening, data extraction and conflict resolution. EH, AD and PAA contributed to visualization. All authors contributed to manuscript review and editing.

## References

Afaya, A., Azongo, T. B., Dzomeku, V. M., Afaya, R. A., Salia, S. M., Adatara, P., Kaba Alhassan, R., Amponsah, A. K., Atakro, C. A., & Adadem, D. (2020). Women’s knowledge and its associated factors regarding optimum utilisation of antenatal care in rural Ghana: A cross-sectional study. PLoS One, 15(7), e0234575.

Akpan, U. B., Asibong, U., Omoronyia, E., Arogundade, K., Agan, T., & Ekott, M. (2020). Severe life threatening pregnancy complications,“near miss” and maternal mortality in a Tertiary Hospital in Southern Nigeria: a retrospective study. Obstetrics and Gynecology International, 2020(1), 3697637.

Asfaha, B. T., Gebremariam, S. H., Gebremariam, G. K., & Weldemariam, A. G. (2022). Knowledge about Obstetric Danger Signs and Related Factors in Reproductive Age Women in the Southeast Zone of Tigray, 2021: A Cross Sectional Study. International journal of reproductive medicine, 2022(1), 7346618.

Boafor, T. (2023). Preventing Maternal Mortality From Hypertensive Disorders Of Pregnancy And Obstetric Haemorrhage: Preventing Maternal Mortality. Postgraduate Medical Journal of Ghana, 12(2), 53–54.

Geller, S. E., Koch, A. R., Garland, C. E., MacDonald, E. J., Storey, F., & Lawton, B. (2018). A global view of severe maternal morbidity: moving beyond maternal mortality. Reproductive health, 15, 31–43.

Gesese, S. S., Mersha, E. A., & Balcha, W. F. (2023). Knowledge of danger signs of pregnancy and health-seeking action among pregnant women: a health facility-based cross-sectional study. Ann Med Surg (Lond), 85(5), 1722–1730. 10.1097/ms9.0000000000000610

Gobran, M. A., Fatah, M. T. A., Ramadan, M. S., Amer, G. A., Rabeh, M. M., Elshafei, R. M., Bosilah, A. H., Khalil, H. H. S., Hassanine, S. T., & Mostafa, M. M. (2021). Educational Program for Pregnant Women Regarding Obstetrics Dangerous Signs in Rural Areas. Open Journal of Obstetrics and Gynecology, 11(5), 529–552.

Hailu, M., Gebremariam, A., & Alemseged, F. (2010). Knowledge about Obstetric Danger Signs among Pregnant Women in Aleta Wondo District, Sidama Zone, Southern Ethiopia. Ethiop J Health Sci, 20(1), 25–32. 10.4314/ejhs.v20i1.69428

Hannola, K., Hoppu, S., Mennander, S., Huhtala, H., Laivuori, H., & Tihtonen, K. (2021). Obstetric early warning system to predict maternal morbidity of pre-eclampsia, postpartum hemorrhage and infection after birth in high-risk women: a prospective cohort study. Midwifery, 99, 103015.

Heazell, A. E., Stacey, T., O’Brien, L. M., Mitchell, E. A., & Warland, J. (2018). Excessive fetal movements are a sign of fetal compromise which merits further examination. Medical Hypotheses, 111, 19–23.

Hibstu, D. T., & Siyoum, Y. D. (2017). Knowledge of obstetric danger signs and associated factors among pregnant women attending antenatal care at health facilities of Yirgacheffe town, Gedeo zone, Southern Ethiopia. Archives of Public Health, 75(1), 35. 10.1186/s13690-017-0203-y

Huang, K., Song, L., Wang, L., Gao, Z., Meng, Y., & Lu, Y. (2014). Advanced abdominal pregnancy: an increasingly challenging clinical concern for obstetricians. International journal of clinical and experimental pathology, 7(9), 5461.

Imani Ramazani, B. E., Mabakutuvangilanga Ntala, S. D., Katuashi Ishoso, D., & Rothan-Tondeur, M. (2023). Knowledge of Obstetric Danger Signs among Pregnant Women in the Eastern Democratic Republic of the Congo. Int J Environ Res Public Health, 20(8). 10.3390/ijerph20085593

Joshi, A., Beyuo, T., Oppong, S. A., Moyer, C. A., & Lawrence, E. R. (2020). Preeclampsia knowledge among postpartum women treated for preeclampsia and eclampsia at Korle Bu Teaching Hospital in Accra, Ghana. BMC Pregnancy Childbirth, 20(1), 625. 10.1186/s12884-020-03316-w

Kiptulon, E. K., Galgalo, D. A., Elmadani, M., Orsolya, M., & Siket, A. U. (2025). Knowledge of obstetric danger signs and birth preparedness and complications readiness among mobile Pokot nomadic pastoralist pregnant women in Tiaty Sub-County, Baringo County- Kenya. BMC pregnancy and childbirth, 25(1), 555. 10.1186/s12884-025-07532-0

Lau, E., Adams, Y. J., Ghiaseddin, R., Sobiech, K., & Ehla, E. E. (2023). Knowledge of postbirth warning signs and postpartum education among women in Ghana. *Journal of Obstetric*, Gynecologic & Neonatal Nursing, 52(4), 309–319.

Negero, M. G., Sibbritt, D., & Dawson, A. (2021). How can human resources for health interventions contribute to sexual, reproductive, maternal, and newborn healthcare quality across the continuum in low-and lower-middle-income countries? A systematic review. Human resources for health, 19, 1–28.

Nkamba, D. M., Wembodinga, G., Bernard, P., Ditekemena, J., & Robert, A. (2021). Awareness of obstetric danger signs among pregnant women in the Democratic Republic of Congo: evidence from a nationwide cross-sectional study. BMC Women’s Health, 21(1), 82. 10.1186/s12905-021-01234-3

Olea-Ramirez, L. M., Leon-Larios, F., & Corrales-Gutierrez, I. (2024). Intervention Strategies to Reduce Maternal Mortality in the Context of the Sustainable Development Goals: A Scoping Review. Women, 4(4), 387–405.

Oni, O. O., Bello, O. O., & Ajayi, A.-O. A. (2025). Determinants of adequate knowledge of postpartum warning signs and complications among parturients in Ibadan, Nigeria: a cross sectional study. BMC pregnancy and childbirth, 25(1), 894. 10.1186/s12884-025-08058-1

Padilla, C., Markwei, M., Easter, S. R., Fox, K. A., Shamshirsaz, A. A., & Foley, M. R. (2021). Critical care in obstetrics: a strategy for addressing maternal mortality. American journal of obstetrics and gynecology, 224(6), 567–573.

Peprah, O. M., Tumu, D., Atibila, F., Ewusie, T. B., & Prempeh, C. A. (2025). Knowledge and associated factors of obstetric danger signs among pregnant women in northern Ghana: a health facility based cross-sectional study. South African Journal of Obstetrics and Gynaecology.

Phanice, O. K., & Zachary, M. O. (2018). Knowledge of obstetric danger signs among pregnant women attending antenatal care clinic at health facilities within Bureti Sub-County of Kericho County, Kenya. Research in Obstetrics and Gynecology, 6(1), 16–21.

Subayi, T. (2023). Where You Live Can Kill You: Key Factors that Contribute to Maternal Morbidity in Resource-Scarce Areas Case Western Reserve University].

Suglo, S., & Siakwa, M. (2016). Knowledge and practices on birth preparedness among expectant mothers seeking antenatal care at the Tamale Teaching Hospital, Ghana. Int J Res, 8(1), 2307–2083.

Sumankuuro, J., Domapielle, M., & Derbile, E. (2022). The what’s, where’s and why’s of miscarriage: evidence from the 2017 Ghana Maternal Health Survey. Public Health, 213, 34–46.

Tamang, S. T., Dorji, T., Yoezer, S., Phuntsho, T., & Dorji, P. (2021). Knowledge and understanding of obstetric danger signs among pregnant women attending the antenatal clinic at the National Referral Hospital in Thimphu, Bhutan: a cross-sectional study. BMC pregnancy and childbirth, 21, 1–9.

Uwiringiyimana, E., Manirambona, E., Byiringiro, S., Nsanzimana, A., Uhawenayo, N., Ufitinema, P., Bayizere, J., Moreland, P. J., Meharry, P., & Ntasumbumuyange, D. (2022). Pregnant women’s knowledge of obstetrical danger signs: A cross-sectional survey in Kigali, Rwanda. PLOS Glob Public Health, 2(11), e0001084. 10.1371/journal.pgph.0001084

Vidler, M., Kinshella, M.-L. W., Sevene, E., Lewis, G., von Dadelszen, P., Bhutta, Z., Sevene, E., Macete, E., Munguambe, K., Sacoor, C., Vala, A., Boene, H., Amose, F., Pires, R., Nhamirre, Z., Macamo, M., Chiaú, R., Matavele, A., Vilanculo, F., … the, C. W. G. (2023). Transitioning from the “Three Delays” to a focus on continuity of care: a qualitative analysis of maternal deaths in rural Pakistan and Mozambique. BMC pregnancy and childbirth, 23(1), 748. 10.1186/s12884-023-06055-w

