## supplemental list of tables for "Determinants Of Knowledge And Awareness Of Obstetric Danger Signs Among Pregnant Women In Kumasi, Ghana: A Cross-Sectional Study"

**Table 4.1: Sociodemographic Characteristics of study participants**

| **Variable** | **Frequency (n=200)** | **Percentage (%)** |
| --- | --- | --- |
| **Age Group (years)** |  |  |
| 18–25 | 61 | 30.5 |
| 26–30 | 71 | 35.5 |
| 31–45 | 68 | 34 |
| **Marital Status** |  |  |
| Single | 70 | 35 |
| Married | 119 | 59.5 |
| Widowed | 4 | 2 |
| Divorced | 7 | 3.5 |
| **Participant's Education** |  |  |
| No formal education | 16 | 8 |
| Primary education | 20 | 10 |
| Secondary education | 77 | 38.5 |
| Tertiary education | 87 | 43.5 |
| **Partner's Education** |  |  |
| No formal education | 15 | 7.5 |
| Primary education | 4 | 2 |
| Secondary education | 69 | 34.5 |
| Tertiary education | 112 | 56 |
| **Current Occupation** |  |  |
| Unemployed | 50 | 25 |
| Student | 26 | 13 |
| Self-employed | 51 | 25.5 |
| Private sector | 32 | 16 |
| Public sector | 41 | 20.5 |
| **Household Monthly Income (GHS)** |  |  |
| <500 | 4 | 2 |
| 500–999 | 19 | 9.5 |
| 1000–1999 | 44 | 22 |
| 2000–2999 | 72 | 36 |
| ≥3000 | 61 | 30.5 |
| **Number of Children** |  |  |
| 0 | 58 | 29 |
| 1–3 | 115 | 57.5 |
| 4–6 | 27 | 13.5 |
| **Religion** |  |  |
| Christianity | 157 | 78.5 |
| Islam | 43 | 21.5 |
| **Residence** |  |  |
| Urban | 198 | 99 |
| Rural | 2 | 1 |
| **Ethnicity** |  |  |
| Akan | 171 | 85.5 |
| Fante | 5 | 2.5 |
| Ewe | 11 | 5.5 |
| Ga | 3 | 1.5 |
| Kusaasi | 2 | 1 |
| Dagomba | 3 | 1.5 |
| Others | 5 | 2.5 |

Data presented as frequency and percentage.

**Table 4.2: Obstetric Characteristics of study participants**

| Variable | Frequency (n=200) | Percentage (%) |
| --- | --- | --- |
| **How many times have you been pregnant (including the current pregnancy)?** |  |  |
| 1 | 45 | 22.5 |
| 1-4 | 124 | 62 |
| > 4 | 31 | 15.5 |
| **Trimester** |  |  |
| First | 52 | 26 |
| Second | 109 | 54.5 |
| Third | 39 | 19.5 |
| **How many times have you given birth to a baby who was born alive (excluding the current pregnancy)?** |  |  |
| 0 | 57 | 28.5 |
| 1-3 | 118 | 59 |
| > 3 | 25 | 12.5 |
| **Complication in previous pregnancy** |  |  |
| No complication | 151 | 75.5 |
| Miscarriage | 24 | 12 |
| Preeclampsia | 10 | 5 |
| Preterm labor | 9 | 4.5 |
| Gestational diabetes | 8 | 4 |
| Abortion | 5 | 2.5 |
| Stillbirth | 2 | 1 |
| Vaginal bleeding | 1 | 0.5 |
| Severe bleeding after birth. Emergency c/s done to correct bleeding | 1 | 0.5 |
| Hyperemesis gravidarium | 1 | 0.5 |
| Complications in current pregnancy |  |  |
| No complications | 174 | 87 |
| Pregnancy-induced hypertension | 6 | 3 |
| Swollen extremities | 5 | 2.5 |
| Abdominal pain | 3 | 1.5 |
| Back pain | 3 | 1.5 |
| Vaginal discharge with odor or itching | 2 | 1 |
| Gestational diabetes | 3 | 1.5 |
| **How many times have you visited the antenatal clinic during this pregnancy?** |  |  |
| 1-3 | 93 | 46.5 |
| 4-6 | 91 | 45.5 |
| 7-9 | 16 | 8 |

Data presented as frequency and percentage

**Table 4.3: Obstetric danger signs identified in the antepartum phase**

| Variable | Frequency (n=200) | Percent (%) |
| --- | --- | --- |
| Severe vaginal bleeding | 194 | 97 |
| Swollen hands, face, and/or feet | 109 | 54.5 |
| Severe pelvic and abdominal pain | 143 | 71.5 |
| No/reduced fetal movement | 162 | 81 |
| Water breaks without labor | 154 | 77 |
| Convulsion | 61 | 30.5 |
| Blurred vision | 71 | 35.5 |
| Severe headache | 114 | 57 |
| Loss of consciousness | 104 | 52 |
| Hyperemesis gravidarum | 2 | 1 |
| High fever | 1 | 0.5 |

Data presented as frequency and percentage

**Table 4.4: Obstetric danger signs identified in the intrapartum phase**

| **Variable** | **Frequency (n=200)** | **Percentage (%)** |
| --- | --- | --- |
| Retained placenta | 190 | 95 |
| Severe vaginal bleeding | 170 | 85 |
| Loss of consciousness | 78 | 39 |
| Labor lasting more than 12 hours | 136 | 68 |
| Convulsion | 31 | 15.5 |
| Baby’s hand/feet come first | 111 | 55.5 |
| Cord comes first | 114 | 57 |
| Difficulty breathing | 50 | 25 |

Data presented as frequency and percentage

**Table 4.5: Obstetric danger signs identified in the postpartum phase**

| **Variable** | **Frequency (n=200)** | **Percent (%)** |
| --- | --- | --- |
| High fever | 85 | 42.5 |
| Severe vaginal bleeding | 184 | 92 |
| Loss of consciousness | 1 | 0.5 |
| Swollen hands, face, and/or feet | 73 | 36.5 |
| Convulsion | 7 | 3.5 |
| Foul-smelling lochia | 68 | 34 |
| Difficulty breathing | 19 | 9.5 |
| Severe lower abdominal pain | 2 | 1 |

Data presented as frequency and percentage

**Table 4.6: Association between sociodemographic variables and good knowledge of obstetric danger signs**

| **Variable** | **Total (n = 200)** | **Poor Knowledge (n = 118)** | **Good Knowledge (n = 82)** | **p value** |
| --- | --- | --- | --- | --- |
| Age Group |  |  |  | **< 0.001** |
| 18–25 | 61 (30.5) | 45 (38.1) | 16 (19.5) |  |
| 26–30 | 71 (35.5) | 45 (38.1) | 26 (31.7) |  |
| 31–45 | 68 (34.0) | 28 (23.7) | 40 (48.8) |  |
| **Marital Status** |  |  |  | **0.014** |
| Single | 70 (35.0) | 52 (44.1) | 18 (22.0) |  |
| Married | 119 (59.5) | 61 (51.7) | 58 (70.7) |  |
| Widowed | 4 (2.0) | 2 (1.7) | 2 (2.4) |  |
| Divorced | 7 (3.5) | 3 (2.5) | 4 (4.9) |  |
| **Level of Education** |  |  |  | **0.001** |
| No formal education | 16 (8.0) | 4 (3.4) | 12 (14.6) |  |
| Primary education | 20 (10.0) | 18 (15.3) | 2 (2.4) |  |
| Secondary education | 77 (38.5) | 46 (39.0) | 31 (37.8) |  |
| Tertiary education | 87 (43.5) | 50 (42.4) | 37 (45.1) |  |
| **Partner’s Level of Education** |  |  |  | 0.888 |
| No formal education | 15 (7.5) | 9 (7.6) | 6 (7.3) |  |
| Primary education | 4 (2.0) | 3 (2.5) | 1 (1.2) |  |
| Secondary education | 69 (34.5) | 39 (33.1) | 30 (36.6) |  |
| Tertiary education | 112 (56.0) | 67 (56.8) | 45 (54.9) |  |
| **Occupation** |  |  |  | **0.016** |
| Unemployed | 50 (25.0) | 35 (29.7) | 15 (18.3) |  |
| Student | 26 (13.0) | 21 (17.8) | 5 (6.1) |  |
| Self-employed | 51 (25.5) | 26 (22.0) | 25 (30.5) |  |
| Private sector | 32 (16.0) | 17 (14.4) | 15 (18.3) |  |
| Public sector | 41 (20.5) | 19 (16.1) | 22 (26.8) |  |
| **Monthly Household Income** |  |  |  | **< 0.001** |
| < GHS 500 | 4 (2.0) | 3 (2.5) | 1 (1.2) |  |
| GHS 500–999 | 19 (9.5) | 19 (16.1) | 0 (0.0) |  |
| GHS 1000–1999 | 44 (22.0) | 28 (23.7) | 16 (19.5) |  |
| GHS 2000–2999 | 72 (36.0) | 39 (33.1) | 33 (40.2) |  |
| ≥ GHS 3000 | 61 (30.5) | 29 (24.6) | 32 (39.0) |  |
| **Number of Children** |  |  |  | **< 0.001** |
| 0 | 58 (29.0) | 48 (40.7) | 10 (12.2) |  |
| 1–3 | 115 (57.5) | 60 (50.8) | 55 (67.1) |  |
| 4–6 | 27 (13.5) | 10 (8.5) | 17 (20.7) |  |
| **Religion** |  |  |  | 0.407 |
| Christianity | 157 (78.5) | 95 (80.5) | 62 (75.6) |  |
| Islam | 43 (21.5) | 23 (19.5) | 20 (24.4) |  |
| **Residence** |  |  |  | 0.795 |
| Urban | 81 (98.8) | 198 (99.0) | 117 (99.2) |  |
| Rural | 1 (1.2) | 2 (1.0) | 1 (0.8) |  |
| **Ethnicity** |  |  |  | 0.176 |
| Akan | 71 (86.6) | 171 (85.5) | 100 (84.7) |  |
| Fante | 0 (0.0) | 5 (2.5) | 5 (4.2) |  |
| Ewe | 5 (6.1) | 11 (5.5) | 6 (5.1) |  |
| Ga | 3 (3.7) | 3 (1.5) | 0 (0.0) |  |
| Kusaasi | 1 (1.2) | 2 (1.0) | 1 (0.8) |  |
| Dagomba | 1 (1.2) | 3 (1.5) | 2 (1.7) |  |
| Others | 1 (1.2) | 5 (2.5) | 4 (3.4) |  |

Data presented as frequency and percentage, p-values computed by Chi-square, p < 0.05 and bolded means statistically significant

**Table 4.7: Association between sociodemographic variables and good knowledge of obstetric danger signs**

| **Variable** | **Total (n = 200)** | **Poor Knowledge (n = 118)** | **Good Knowledge (n = 82)** | **p value** |
| --- | --- | --- | --- | --- |
| **How many times have you been pregnant (including the current pregnancy)** |  |  |  | **<0.001** |
| 1 | 45 (22.5) | 38 (32.2) | 7 (8.5) |  |
| 2–4 | 124 (62.0) | 71 (60.2) | 53 (64.6) |  |
| > 4 | 31 (15.5) | 9 (7.6) | 22 (26.8) |  |
| **Trimester** |  |  |  | **0.002** |
| First | 52 (26.0) | 41 (34.7) | 11 (13.4) |  |
| Second | 109 (54.5) | 59 (50.0) | 50 (61.0) |  |
| Third | 39 (19.5) | 18 (15.3) | 21 (25.6) |  |
| **Parity** |  |  |  | **<0.001** |
| 0 | 57 (28.5) | 47 (39.8) | 10 (12.2) |  |
| 1–3 | 118 (59.0) | 64 (54.2) | 54 (65.9) |  |
| > 3 | 25 (12.5) | 7 (5.9) | 18 (22.0) |  |
| **Have you experienced any complications in your previous pregnancies** |  |  |  | **0.001** |
| Yes | 49 (24.5) | 19 (16.1) | 30 (36.6) |  |
| No | 151 (75.5) | 99 (83.9) | 52 (63.4) |  |
| **Complications in current pregnancy** |  |  |  | **0.001** |
| No Complications | 174 (87.0) | 111 (94.1) | 63 (76.8) |  |
| Pregnancy-induced hypertension | 6 (3.0) | 0 (0.0) | 6 (7.3) |  |
| Swollen extremities | 9 (4.5) | 3 (2.5) | 6 (7.3) |  |
| Abdominal pain | 3 (1.5) | 3 (2.5) | 0 (0.0) |  |
| Back pain | 3 (1.5) | 0 (0.0) | 3 (3.7) |  |
| Vaginal discharge with odor or itching | 2 (1.0) | 0 (0.0) | 2 (2.4) |  |
| Gestational diabetes | 3 (1.5) | 1 (0.8) | 2 (2.4) |  |
| **ANC Visits during current pregnancy** | |  |  | **0.004** |
| 1–3 | 93 (46.5) | 65 (55.1) | 28 (34.1) |  |
| 4–6 | 91 (45.5) | 48 (40.7) | 43 (52.4) |  |
| 6–9 | 16 (8.0) | 5 (4.2) | 11 (13.4) |  |

Data presented as frequency and percentage, p-values computed by Chi-square, p < 0.05 and bolded means statistically significant

**Table 4.8: Sociodemographic Predictors of good knowledge of obstetric danger signs**

| **Variable** | **Good Knowledge n (=82)** | **cOR (95% CI)** | **p-value** | **aOR (95% CI)** | **p-value** |
| --- | --- | --- | --- | --- | --- |
| **Age Group** |  |  |  |  |  |
| 18–25 (Ref) | 16 (19.5) | 1 | – |  |  |
| 26–30 | 26 (31.7) | 1.63 (0.77–3.43) | 0.203 | 0.56 (0.19–1.62) | 0.284 |
| 31–45 | 40 (48.8) | 4.02 (1.90–8.48) | **< 0.001** | 1.03 (0.35–3.01) | 0.962 |
| **Marital Status** |  |  |  |  |  |
| Single (Ref) | 18 (22.0) | 1 | – |  |  |
| Married | 58 (70.7) | 2.75 (1.44–5.24) | **0.002** | 1.21 (0.54–2.73) | 0.648 |
| Widowed | 2 (2.4) | 2.89 (0.38–22.04) | 0.306 | 0.90 (0.08–10.42) | 0.933 |
| Divorced | 4 (4.9) | 3.85 (0.79–18.89) | **0.096** | 5.07 (0.54–47.25) | 0.154 |
| **Level of Education** |  |  |  |  |  |
| No formal education (Ref) | 12 (14.6) | 1 | – |  |  |
| Primary education | 2 (2.4) | 0.04 (0.006–0.24) | **< 0.001** | 0.06 (0.01–0.46) | **0.007** |
| Secondary education | 31 (37.8) | 0.23 (0.066–0.76) | **0.016** | 0.44 (0.11–1.76) | 0.245 |
| Tertiary education | 37 (45.1) | 0.25 (0.074–0.83) | **0.023** | 0.50 (0.10–2.49) | 0.399 |
| **Occupation** |  |  |  |  |  |
| Unemployed (Ref) | 15 (18.3) | 1 | – |  |  |
| Student | 5 (6.1) | 0.56 (0.18–1.75) | 0.315 | 1.75 (0.34–8.96) | 0.500 |
| Self-employed | 25 (30.5) | 2.24 (0.99–5.08) | 0.053 | 1.64 (0.60–4.52) | 0.337 |
| Private sector | 15 (18.3) | 2.06 (0.82–5.17) | 0.124 | 1.47 (0.51–4.25) | 0.473 |
| Public sector | 22 (26.8) | 2.70 (1.14–6.40) | **0.024** | 1.58 (0.46–5.43) | 0.465 |
| **Monthly Income** |  |  |  |  |  |
| < GHS 500 (Ref) | 1 (1.2) | 1 | – |  |  |
| GHS 500–999 | 0 (0.0) | ~0.00 (NA) | 0.998 | 0.00 (0.00–∞) | 0.998 |
| GHS 1000–1999 | 16 (19.5) | 1.71 (0.16–17.89) | 0.652 | 1.12 (0.09–14.00) | 0.93 |
| GHS 2000–2999 | 33 (40.2) | 2.54 (0.25–25.58) | 0.429 | 1.14 (0.09–14.46) | 0.922 |
| ≥ GHS 3000 | 32 (39.0) | 3.31 (0.33–33.63) | 0.312 | 1.17 (0.09–16.15) | 0.907 |
| **Number of Children** |  |  |  |  |  |
| 0 (Ref) | 10 (12.2) | 1 | – |  |  |
| 1–3 | 55 (67.1) | 4.40 (2.03–9.54) | **< 0.001** | 3.86 (1.31–11.36) | **0.014** |
| 4–6 | 17 (20.7) | 8.16 (2.89–23.01) | **< 0.001** | 6.12 (1.43–26.16) | **0.015** |

cOR: crude odds ratio, aOR: adjusted odds ratio, CI: Confidence interval, p < 0.05 and bolded means statistically significant

**Table 4.9: Obstetric Predictors of good knowledge of obstetric danger signs**

| **Variable** | **Good Knowledge (n = 82)** | **cOR (95% CI)** | **p-value** | **aOR (95% CI)** | **p-value** |
| --- | --- | --- | --- | --- | --- |
| **How many times have you been pregnant (including the current pregnancy)** | | | | | |
| 1 (Ref) | 7 (8.5) | 1 | - | 1 | - |
| 2–4 | 53 (64.6) | 4.05 (1.68–9.78) | **0.002** | 0.82 (0.12–5.84) | 0.843 |
| > 4 | 22 (26.8) | 13.27 (4.34–40.61) | **<0.001** | 0.63 (0.03–13.67) | 0.767 |
| **Trimester** | |  |  |  |  |
| First (Ref) | 11 (13.4) | 1 | - | 1 | - |
| Second | 50 (61.0) | 3.16 (1.47–6.79) | **0.003** | 1.05 (0.33–3.37) | 0.940 |
| Third | 21 (25.6) | 4.35 (1.74–10.87) | **0.002** | 0.70 (0.14–3.49) | 0.661 |
| **Parity** |  |  |  |  |  |
| 0 (Ref) | 10 (12.2) | 1 | - | 1 | - |
| 1–3 | 54 (65.9) | 3.97 (1.83–8.59) | **<0.001** | 4.61 (0.83–25.65) | 0.081 |
| > 3 | 18 (22.0) | 12.09 (3.99–36.61) | **<0.001** | 17.88 (0.83–383.82) | 0.065 |
| **Have you experienced any complications in your previous pregnancies** | | | | | |
| Yes (Ref) | 30 (36.6) | 1 | - | 1 | - |
| No | 52 (63.4) | 0.33 (0.17–0.65) | **0.001** | 0.24 (0.09–0.65) | **0.005** |
| **Complications in current pregnancy** | | | |  |  |
| No Complications (Ref) | 63 (76.8) | 1 | - | 1 | - |
| Pregnancy-induced hypertension | 6 (7.3) | 7.05 (0.77–64.44) | 0.084 | 13.25 (1.10–159.02) | **0.042** |
| Swollen extremities | 6 (7.3) | 1.76 (0.24–12.82) | 0.576 | 1.39 (0.15–12.57) | 0.768 |
| Abdominal pain | 0 (0.0) | 0.00 (0.00–∞) | 0.999 | 0.00 (0.00–∞) | 0.999 |
| Back pain | 3 (3.7) | - | 0.999 | - | 0.999 |
| Vaginal discharge with odor or itching | 2 (2.4) | - | 0.999 | - | 0.999 |
| Gestational diabetes | 2 (2.4) | 3.52 (0.31–39.64) | 0.308 | 0.71 (0.05–9.87) | 0.801 |
| **ANC Visits during current pregnancy** | | | |  |  |
| 1–3 (Ref) | 28 (34.1) | 1 | - | 1 | - |
| 4–6 | 43 (52.4) | 2.08 (1.14–3.81) | **0.018** | 1.81 (0.70–4.71) | 0.222 |
| 6–9 | 11 (13.4) | 5.11 (1.62–16.07) | **0.005** | 8.98 (1.44–56.12) | **0.019** |

cOR: crude odds ratio, aOR: adjusted odds ratio, CI: Confidence interval, p < 0.05 and bolded means statistically significant

**Table 4.10: Awareness of Obstetric danger signs**

| **Statement** | **Yes n (%)** | **No n (%)** |
| --- | --- | --- |
| Recognizing warning signs is important for your health and your baby’s health | 173 (86.5) | 27 (13.5) |
| Aware that danger signs require immediate medical attention | 158 (79.0) | 42 (21.0) |
| Awareness can help prevent serious complications or death | 147 (73.5) | 53 (26.5) |
| Important to have an emergency plan during pregnancy | 181 (90.5) | 19 (9.5) |
| Early detection and treatment of complications improves delivery outcomes | 153 (76.5) | 47 (23.5) |
| Seeking help for serious symptoms can protect your health | 154 (77.0) | 46 (23.0) |

Data presented as frequency and percentage
